# Heterogeneity in and correlation between host transmissibility and susceptibility can greatly impact epidemic dynamics

**DOI:** 10.1101/2024.12.10.24318805

**Authors:** Beth M. Tuschhoff, David A. Kennedy

## Abstract

While it is well established that host heterogeneity in transmission and host heterogeneity in susceptibility each individually impact disease dynamics in characteristic ways, it is generally unknown how disease dynamics are impacted when both types of heterogeneity are simultaneously present. Here we explore this question. We first conducted a systematic review of published studies from which we determined that the effects of correlations have been drastically understudied. We then filled in the knowledge gaps by developing and analyzing a stochastic, individual-based SIR model that includes both heterogeneity in transmission and susceptibility and flexibly allows for positive or negative correlations between transmissibility and susceptibility. We found that in comparison to the uncorrelated case, positive correlations result in major epidemics that are larger, faster, and more likely, whereas negative correlations result in major epidemics that are smaller and less likely. We additionally found that, counter to the conventional wisdom that heterogeneity in susceptibility always reduces outbreak size, heterogeneity in susceptibility can lead to major epidemics that are larger and more likely than the homogeneous case when correlations between transmissibility and susceptibility are positive, but this effect only arises at small to moderate *R*_0_. Moreover, positive correlations can frequently lead to major epidemics even with subcritical *R*_0_. To illustrate the potential importance of heterogeneity and correlations, we developed an SEIR model to describe mpox disease dynamics in New York City, demonstrating that the dynamics of a 2022 outbreak can be reasonably well explained by the presence of positive correlations between susceptibility and transmissibility. Ultimately, we show that correlations between transmissibility and susceptibility profoundly impact disease dynamics.

**Highlights:**

- Systematic review finds that effects of correlations on epidemics are understudied
- Positive correlations lead to larger, faster, more likely epidemics
- Negative correlations lead to smaller, less likely epidemics
- Positive correlations allow for major epidemics even with subcritical *R*_0_
- Mpox outbreak dynamics in New York City were consistent with positive correlations

## 1. Introduction

Some epidemics are fast and explosive while others are slow and meandering. These differences could arise in many ways, including from differences in factors such as *R*_0_ values or transmission rates (Anderson and May, 1991; Heffernan et al., 2005). Here we explore whether heterogeneity in and correlations between host transmissibility and susceptibility can also drive changes in disease dynamics.

It is well established that host heterogeneity can have large impacts on disease dynamics (Dwyer et al., 1997; Lloyd-Smith et al., 2005; Gomes et al., 2014; Langwig et al., 2017; Gomes et al., 2022). Compared to the case without heterogeneity, heterogeneity in individuals’ likelihoods of transmitting a pathogen once infected, hereafter referred to as “heterogeneity in transmission”, results in epidemics that are rarer and more likely to go extinct but initially more explosive when they do take off (Lloyd-Smith et al., 2005). Likewise, heterogeneity in individuals’ likelihoods of becoming infected, hereafter referred to as “heterogeneity in susceptibility”, results in a lower peak number of cases and a smaller final epidemic size (Aguas et al., 2020; Gomes et al., 2022; Montalbán et al., 2022). But what happens when a population has heterogeneity in both transmission and susceptibility? Moreover, how do individual-level correlations in transmissibility and susceptibility affect disease dynamics? Here we explore the effects of positive and negative correlations on disease dynamics, first by conducting a systematic literature review and second by developing a finite population model that flexibly allows for positive or negative correlations between host transmissibility and host susceptibility.

Positive and negative correlations between transmissibility and susceptibility can arise in many ways. Focusing first on positive correlations, one way that positive correlations can arise is when individuals are heterogeneous in their contact rates. Individuals with more contacts, such as individuals with careers that require public interface and social interactions (Baker et al., 2020), will presumably be at higher risk for infection and be more likely to transmit a pathogen once infected. Likewise, individuals may vary in their risk taking behavior in ways that make them both more susceptible to infection and more transmissible given infection. For instance, individuals who engage in high risk sexual behaviors are more likely to both contract and transmit sexually transmitted infections (Ericksen and Trocki, 1992). Coinfection may also lead to positive correlations betweeen susceptibility and transmissibility when primary infections weaken immune responses (Galvin and Cohen, 2004; Zélé et al., 2018; Karvonen et al., 2019). Likewise, individuals that are immunocompromised for other reasons may be at greater risk for infection and have increased shedding, leading to more transmission (Permar et al., 2006; Memoli et al., 2014).

Although perhaps less intuitive, negative correlations can likewise arise in multiple ways. For example, when severe disease results in an individual changing their behavior such that they reduce their contact rates, immunocompromised individuals would likely be at greater risk of infection but less likely to transmit infection once it occurs (Collins et al., 2020; Li et al., 2021; Kennedy, 2023). Similarly, if individuals that are resistant to infection are also more likely than average to develop asymptomatic infections, they may end up transmitting at higher rates than average because they are unaware of their infection status (Kalajdzievska and Li, 2011; Medzhitov et al., 2012). Certain types of public health interventions could additionally create negative correlations between susceptibility and transmissibility. For instance, vaccination targeted towards people with highly social jobs could lower their overall susceptibility to infection, but if infected, these individuals may still transmit at high rates (Nunner et al., 2022; Park et al., 2022). Similarly, during disease outbreaks, front line workers may be at high risk for infection due to high rates of pathogen exposure, but may also be closely monitored and quarantined at the first signs of illness, reducing opportunities for the onward transmission of infection (Bielicki et al., 2020).

There has been substantial work conducted on understanding the role of host heterogeneity in transmission or susceptibility on disease dynamics. In this paper, we explored the role of interactions between these factors. We first conducted a systematic review of published studies that investigated the effects of correlations between transmissibility and susceptibility on disease dynamics to identify knowledge gaps in this topic area. To fill these knowledge gaps, we then developed a stochastic, individual-based SIR model that includes both heterogeneity in transmission and susceptibility and that flexibly allows for positive, negative, or zero correlations between transmissibility and susceptibility. After exploring the effects of correlations with this theoretical model, we applied an expanded SEIR version of it to mpox. Mpox is spread mainly through close contact, presumably resulting in a strong positive correlation between susceptibility and transmissibility (Laurenson-Schafer et al., 2023). We show that the explosive dynamics of the 2022 mpox epidemic, followed by its rapid decline but long term persistence at low prevalence, is consistent with what would be expected if there were strong positive correlations between susceptibility and transmissibility. The 2022 mpox epidemic may thus provide a real-world example of the importance of correlations on understanding infectious disease dynamics.

## 2. Definitions of the basic and effective reproductive numbers

Infectious disease dynamics are frequently modeled using a series of ordinary differential equations that track the infection status of hosts. Such models are frequently termed SIR or SEIR models which allow hosts to transition through compartments of susceptible, infectious, and removed (with an added exposed-but-not-yet-infectious class in the SEIR model). The simplest verision of the SIR framework is

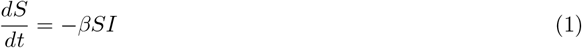

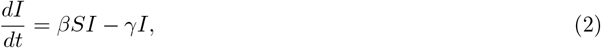

where *S* is the number of susceptible individuals, *I* is the number of infected individuals, and the removed individuals are not explicitly tracked. In the above model, *β* is the transmission rate and *γ* is the recovery rate. A key result from previous analyses of these types of models is that there is a threshold value above which outbreaks can occur 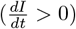 and below which outbreaks cannot occur 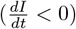. This threshold, in a fully naive host population, is termed *R*_0_ or the basic reproductive number. *R*_0_ is a valuable parameter for inferring disease dynamics and responding to infectious disease threats (Shaw and Kennedy, 2021). Using its standard definition, *R*_0_ is the expected number of new infections caused by an initial infection in a fully susceptible population (Anderson and May, 1991). For the dynamics described above, 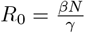 where *N* is the total host population size.

An assumption of the above model framework is that all individuals are equally susceptible to infection and equally transmissible. If this were not the case, we would need to modify the model to account for this variability. In particular, the rate of new infections would no longer be *βSI*, but would instead be described by the force of infection created by each infected host *i* (*λ*_*i*_) multiplied by the risk of infection experienced by each susceptible host *j* (*r*_*j*_) and summed over all combinations of *i* and *j*. Thus the rate of new infections can be described as 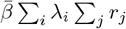 where 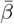 is a scaling constant.

Adding heterogeneity in this way into the model creates a new challenge. Despite the seemingly clear definition of *R*_0_ described above (the number of new infections created by an initial infection), this definition becomes ambiguous in the presence of heterogeneity, because the number of new infections created will depend on which individual is the first one to become infected. To illustrate, consider that *p*_*i*_ is the probability that a particular individual *i* is the first to be infected. Then, we can unambiguously define *R*_0_ as

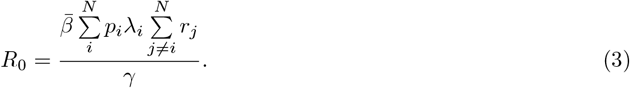

Keeping in mind that 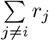 is equal to 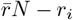 where 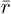 is the mean risk, we can rewrite Eq 3 as

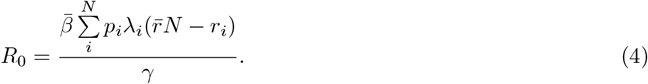

When 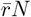 is large relative to the maximum value of *r*_*i*_ (which will be true at large *N*), this can be further simplified to

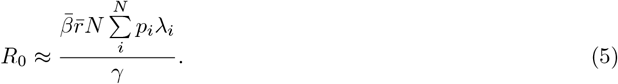

Notably, Eq 5 illustrates that the value of *R*_0_ depends on how likely each individual is to be the initial infection (i.e., the distribution of *p*_*i*_). When each individual is equally likely to be the first one infected (i.e., 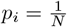 for all *i*),

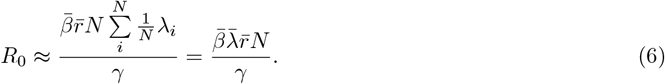

However, when the probability of being the first infection is proportional to each individual’s relative susceptibility 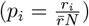

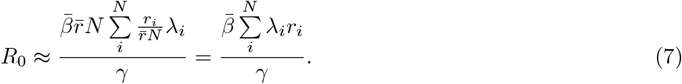

Eqs 6 and 7 thus provide two different definitions of *R*_0_ that need not be similar. The first definition (Eq 6) states that *R*_0_ scales with the product of the average susceptibility and average transmissibility, and the second definition (Eq 7) states that *R*_0_ scales with average of the product of each individual’s susceptibility and transmissibility. Notably, these differences arise due to differences in *p*_*i*_ which would be generally unknowable prior to the initiation of an outbreak. It is worth pointing out that these are simply two definitions, but any distribution of *p*_*i*_ could be used to create a definition for *R*_0_. In this manuscript, we primarily use the definition of *R*_0_ where every individual is equally likely to be the first infected (Eq 6), and we explicitly state when this is not the case.

The effective reproductive number *R*_*e*_, defined as the expected number of new infections caused by an average infectious individual in a population at a current point in time, can be used analogously to *R*_0_ to determine whether an outbreak is currently growing or shrinking. However, *R*_*e*_ does not have the same ambiguity in definition as *R*_0_. This is because once the epidemic begins, each individual has a particular status as infected or not meaning that the parameter *p*_*i*_ is no longer present. Thus, *R*_*e*_ can be unambiguously defined as

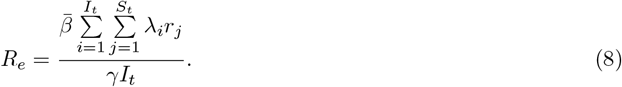

As can be seen in Eq 8 above, the value of *R*_*e*_ depends on the product of *λ*_*i*_ and *r*_*j*_ for all *i* and *j* and therefore the growth rate of an epidemic can change greatly depending on whether the individuals most likely to become infected at any point in time are also the most infectious or the least infectious. This is the conceptual basis behind the idea that correlations between susceptibility and transmissibility can drastically alter disease dynamics.

## 3. Systematic review

To explore the work that has previously been published on the effects of correlations between transmissibility and susceptibility on disease dynamics, we conducted a systematic review.

### 3.1. Systematic review methods

Our systematic review was conducted following the PRISMA guidelines (Fig 1; Page et al. 2021). We searched Google Scholar on January 30, 2024 using the following keywords:

**Figure 1:**
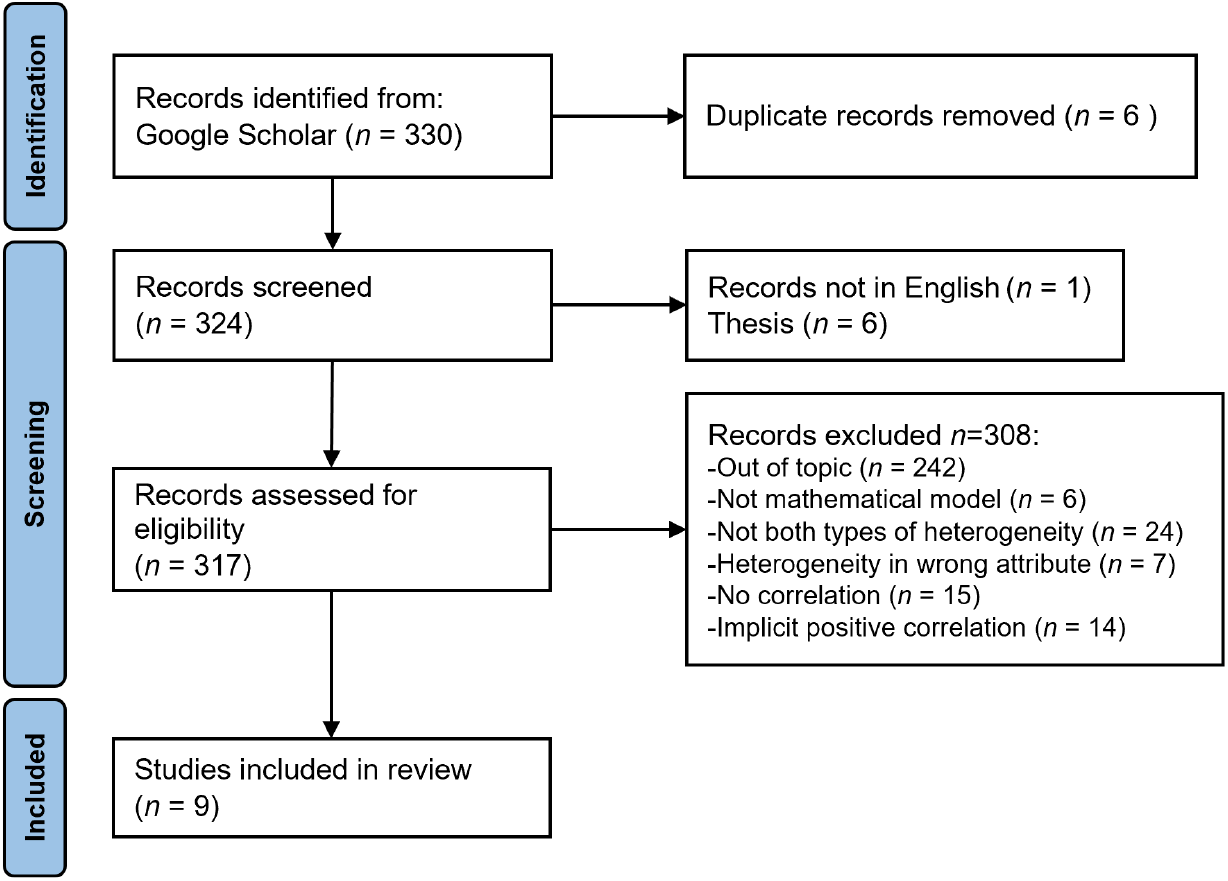
PRISMA systematic review framework.

(“heterogeneity in susceptibility” OR “variation in susceptibility” OR “heterogeneous susceptibility” OR “differential susceptibility”) AND (“heterogeneity in transmission” OR “heterogeneity in infectivity” OR “heterogeneous infectivity” OR “variation in infectivity” OR “variation in infectiousness” OR “differential infectivity”) AND (correlation) AND (infectious disease).

We screened an initial 330 published papers by implementing the following selection criteria:

1. The study must assess a mathematical model.
2. The model must include both heterogeneity in transmission, in terms of an individual’s likelihood of spreading infection, and heterogeneity in susceptibility, in terms of an individual’s likelihood of becoming infected.
3. The model must explore the effect of a correlation between transmissibility and susceptibility on infection dynamics.
4. The model must not implicitly assume a perfect positive correlation (e.g., an undirected network model).

A total of 9 papers fulfilled the selection criteria (Cardell and Kanouse, 1989; Becker and Marschner, 1990; Daley et al., 2000; Andreasen, 2011; Clancy and Pearce, 2013; Hickson and Roberts, 2014; Tkachenko et al., 2021; Kawagoe et al., 2021; Allard et al., 2023). From these selected papers, we recorded the author affiliations, the model structure, the assumptions, and what the model found with regard to the effects of positive or negative correlations between susceptibility and transmissibility on the probability of a major epidemic, the peak size, the peak time, the final epidemic size, and the time to the *j*th infection. For two of the papers, two different sets of assumptions were explored in each. We thus included each of these model versions as separate models in our analyses. This gave us a total of 11 models across the 9 papers that explored how heterogeneity in susceptibility and transmission affect disease dynamics in situations where susceptibility and transmissibility are positively or negatively correlated.

### 3.2. Systematic review results

The 11 models from our 9 selected studies were built using various model structures and assumptions as presented in Fig 2. Eight of 11 models used an SIR-type model structure, 7 of 11 models were deterministic, and 4 of the 11 models assumed that host variation fell into discrete categories. The specific types of distributions used to model heterogeneity differed between models including gamma distributions, unnamed distributions where the authors set their own values, other distributions including lognormal or uniform, and a combination of gamma and another distribution. Ten out of 11 models assumed that the population was well-mixed with the exception being a directed network model. Ten out of 11 models also assumed that the size of the susceptible population changed over time due to individuals transitioning from susceptible to infectious, with the exception being a branching process model. When exploring the effects of correlations betweeen susceptibility and transmissibility on disease dynamics, 4 of the 11 models kept the mean susceptibility and mean transmissibility constant (i.e., our preferred definition of *R*_0_: Eq 6) and 5 of 11 kept the mean product of susceptibility and transmissibility constant (i.e., an alternative definition of *R*_0_: Eq 7). The remaining 2 models (“Other”) respectively kept the median product of susceptibility and transmissibility constant or kept the growth rate for the first month of the epidemic constant. We additionally found that the papers’ authors came from various disciplines but were primarily associated with departments in the fields of physics, math, and biophysics (Fig 3).

**Figure 2:**
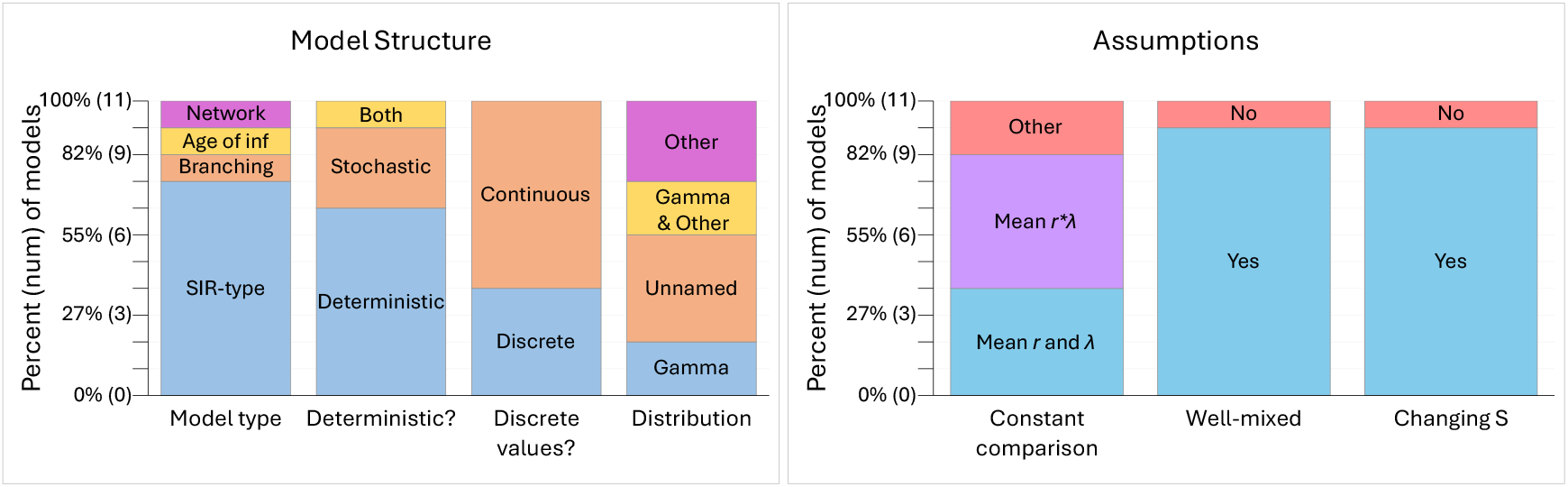
Models were built using various model structures and assumptions. These plots show the number of models that were built under each structure and assumption. For model structure, the model type is either an SIR-type (SIR, SI, etc.), a branching process, an age of infection model, or a directed network. Models were set up as deterministic, stochastic, or both to explore different results. The distributions used to model the values for susceptibility and transmissibility were either discrete or continuous and the type of distribution explored in each study was a gamma distribution, an unnamed distribution where authors set their own values, a gamma distribution plus another distribution, or solely another type of distribution like a lognormal or uniform distribution. For assumptions, models compare the effects of correlation to the uncorrelated and homogeneous cases by keeping the mean susceptibility and mean transmissibility constant (i.e., *R*_0_ defined as in Eq 6; “Mean *r* and *λ*”), keeping the mean product of susceptibility and transmissibility constant (i.e., *R*_0_ defined as in Eq 7; “Mean *r* * *λ*”), or by keeping the growth rate for the first month of the epidemic or the median product of susceptibility and transmissibility constant (“Other”). Most models assume a well-mixed population and a changing susceptible population size over time.

**Figure 3:**
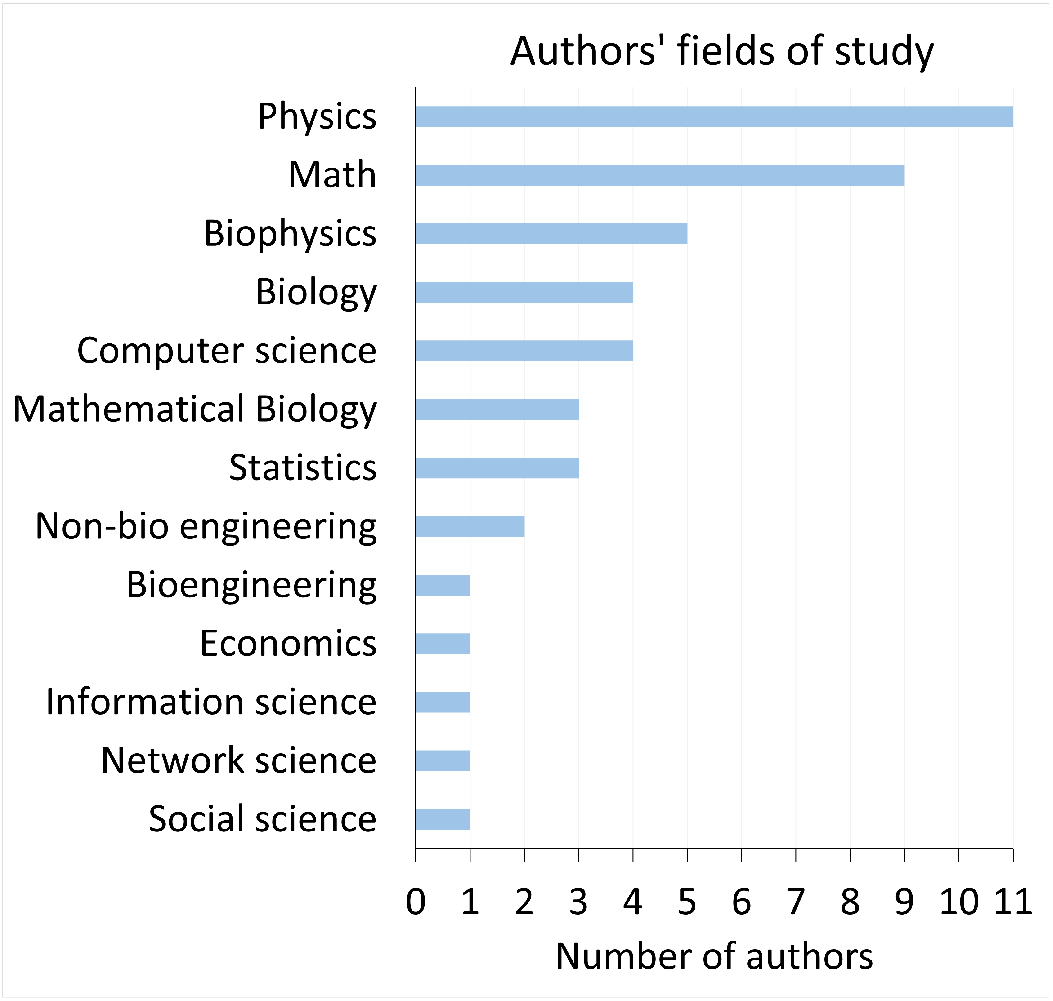
Authors of the selected studies in our systematic review tend to be associated with quantitative fields, especially physics, math, and biophysics. This plot shows the number of authors affiliated with each of the different fields of study represented in the selected studies. Authors’ affiliations were identified by their department or research group and focus at the time the study was published. While there are 31 authors across the studies, some authors are represented multiple times as they are affiliated with multiple fields.

Among the 11 models, the most studied attributes were the final epidemic size and the peak time (Fig 4). The probability of a major epidemic, peak size, and time to the *j*th infection were not as commonly studied. These attributes were only explored by at most two models, and often the models disagreed on the results. In addition, the majority of models explored the effects of a positive correlation between susceptibility and transmissibility but not the effects of a negative correlation. Our review further showed that almost all of the models studied the effects of correlations relative to the case where populations were homogeneous rather than where populations were heterogeneous but susceptibility and transmissibility were uncorrelated. The lone exceptions were two models that investigated the impact of positive correlations on the final epidemic size. Thus, no models explored how negative correlations affected disease dynamics relative to populations with the same heterogeneity, but in which susceptibility and transmissibility were uncorrelated.

**Figure 4:**
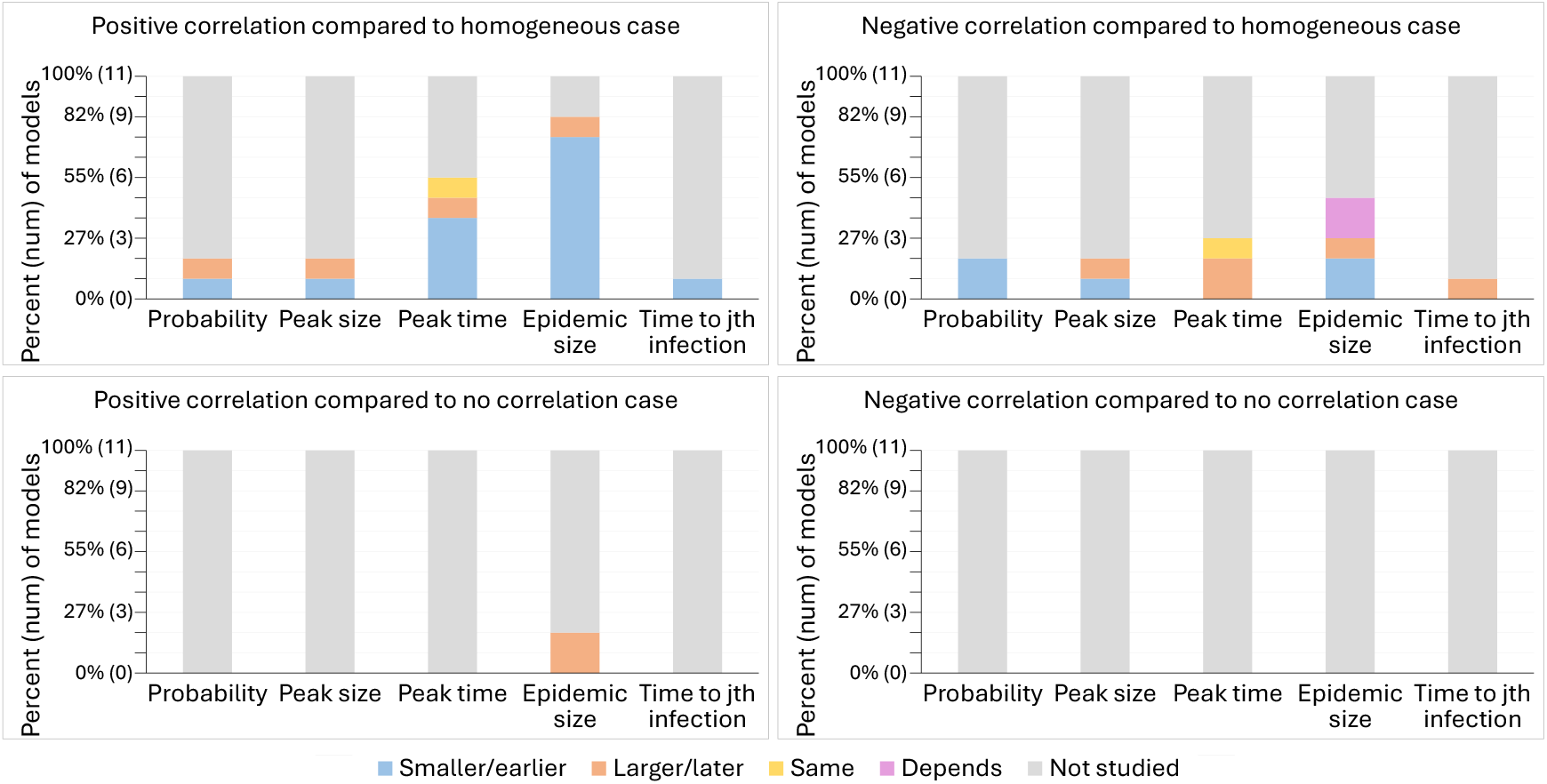
Models tend to disagree about the effects of correlation, and many epidemic measures are understudied, especially comparisons to the no correlation case. These plots show the effects of positive and negative correlations between heterogeneity in transmission and susceptibility on the probability of a major epidemic, peak size, peak time, final epidemic size, and time to the *j*th infection in comparison to disease dynamics with homogeneity or no correlation according to 11 models from the 9 studies included in the systematic review. The effect of correlation is classified for each model and measure as resulting in an attribute that is smaller/earlier (blue), larger/later (orange), the same (yellow), dependent on the levels of heterogeneity (purple), or not studied (gray) in comparison to the homogeneous or no correlation case.

While most models seemed to agree that positive correlations lead to a smaller final epidemic size and earlier peak time than dynamics with homogeneous transmission and susceptibility, we did not find total agreement. Two of 2 models agreed that positive correlations result in larger final epidemic sizes compared to the no correlation case, and 2 of 2 models agreed that negative correlations lead to a smaller probability of a major epidemic in comparison to the homogeneous case. Nevertheless, we found substantial disagreement among many of the models in how correlations impact other epidemic features relative to the homogeneous case, including whether either type of correlation leads to a smaller or larger peak size, whether positive correlations lead to a smaller or larger probability of a major epidemic, whether negative correlations delay the timing of the peak, and whether negative correlations lead to a smaller, larger, or similar final epidemic size. We suspect the disagreement between models is due to differing model structures, different assumptions, different levels of heterogeneity, and different values of *R*_0_. For example, the model that found that positive correlations result in larger final epidemic sizes kept the median of individuals’ *R*_0_ values constant for comparison between the heterogeneous and homogeneous cases, whereas all of the models that kept *R*_0_ constant, regardless of how they defined *R*_0_ (Eq 6 or 7), did not find this effect. Likewise, two models found that compared to the homogeneous case, the effects of a negative correlation on final epidemic size depended on the levels of heterogeneity.

Taken together, our systematic review reveals that the effects of correlation on disease dynamics were understudied by previous models. In particular, the effects of negative correlations and the comparison of dynamics to the uncorrelated case need to be studied. Additionally, the specific attributes of the probability of a major epidemic, peak size, and time to the *j*th infection were not as well-studied and should be examined further. Tables containing the full results of the systematic review as associated with each model are in the Supplement (Tables S1 and S2).

## 4. Theoretical model

To fill in the knowledge gaps and ambiguities identified from our systematic review, we generated an individual-based, stochastic SIR model that flexibly allows us to generate populations of hosts that are heterogeneous or not, and in which we can flexibly alter heterogeneity without altering *R*_0_ (Eq 6). Using our model, we generated populations of hosts that have positive, negative or noncorrelated relationships between transmissibility and susceptibility. With this model, we explore the impact that positive and negative correlations have relative to the uncorrelated case and the homogeneous case. We assessed the same epidemic measures as in the systematic review as well as the effective reproductive number *R*_*e*_. Below, we describe how we generated an extension of this model to describe mpox disease dynamics in New York City during a 2022 outbreak. This extension demonstrates the effects that correlations between susceptibility and transmissibility can have on disease dynamics in real systems. Table 1 contains definitions and values for the parameters used in our models.

**Table 1:** Definitions and values of model parameters. Note that some parameters are simply combinations of other parameters, and values for those parameters are not explicitly listed in this table. Dashes indicate parameters that do not exist for that particular model.

| Symbol | Definition | Theoretical Value | Mpox Value |
| --- | --- | --- | --- |
| $N$ | Population size | 1,000 | 70,180 |
| $\lambda_i$ | Force of infection for the $i$th individual | | |
| $\bar{\lambda}$ | Average force of infection | 1 | 1 |
| $r_i$ | Risk of infection for the $i$th individual | | |
| $\bar{r}$ | Average risk of infection | 1 | 1 |
| $p_i$ | Probability of the $i$th individual being the first individual infected | $\frac{1}{N}$ | $\frac{1}{N}$ |
| $\bar{\beta}$ | Scaling constant for the average transmission rate | 0.00008, 0.00011, 0.0003 | 0.00000054 |
| $\beta_{i,j}$ | Transmission rate from individual $i$ to $j$; $\bar{\beta}\lambda_i r_j$ | | |
| $b$ | Rate at which individuals enter the population | - | 70 |
| $\mu$ | Rate at which individuals leave the population | - | 0.000997 |
| $\delta$ | Rate at which individuals transition from exposed to infectious | - | 0.13 |
| $\gamma$ | Recovery rate | 0.1 | 0.071 |
| $R_0$ | Basic reproductive number | 0.8, 1.1, 3 | 0.52 |
| $R_e$ | Effective reproductive number | | |
| $m$ | Parameter describing the distribution of individuals' forces of infection | | |
| $k$ | Parameter describing the distribution of individuals' risks of infection | | |
| $C_t$ | Coefficient of variation in the force of infection; Level of heterogeneity in transmission; $\frac{1}{\sqrt{m}}$ | 0, 0.5, 1, 3 | 3 |
| $C_s$ | Coefficient of variation of the risk of being infected; Level of heterogeneity in susceptibility; $\frac{1}{\sqrt{k}}$ | 0, 0.5, 1, 3 | 3 |
| $\rho$ | Correlation between transmissibility and susceptibility | -1, -0.5, 0, 0.5, 1 | -1, 0, 1 |

### 4.1. Theoretical model methods

For our theoretical model, we start with the basic SIR framework as described in Eqs 1 and 2. We then include heterogeneity in transmission and susceptibility. To do so, we assign each susceptible individual *i* a unique susceptibility risk *r*_*i*_, and each infected individual *j* a unique force of infection *λ*_*j*_. Prior to the initiation of an outbreak, the expected values 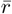 and 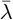 are both normalized to be 1 as described below. Next, we assume that the rate of transmission is multiplicatively determined by these parameters such that the transmission rate from individual *i* to *j* is 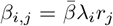, where 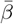 is a constant that scales the average transmission rate, which we use to set *R*_0_ according to Eq 6.

To run our model, we assign each individual *i, i* = 1, …, *N* in the population a relative force of infection *λ*_*i*_ and a relative risk of being infected *r*_*i*_. We set correlation *ρ* between these parameters using a normal copula with marginal gamma distributions where *λ*_*i*_ ~ Gamma(*m*, 1*/m*) and *r*_*i*_ ~ amma(*k*, 1*/k*). We chose to use gamma distributions because they are flexible and have been used to model heterogeneous populations previously (Dwyer et al., 1997; Langwig et al., 2017). Note that we facilitated comparisons across different levels of heterogeneity by setting the mean risk 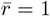 and mean force of infection 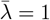, keeping *R*_0_ constant according to our working definition of *R*_0_ (Eq 6). We describe the level of heterogeneity in transmission as *C*_*t*_, which is the coefficient of variation in the force of infection across all potential hosts in the population, where the coefficient of variation is defined as the standard deviation divided by the mean. Being gamma distributed, this equates to 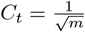. Similarly, we describe the level of heterogeneity in susceptibility using the coefficient of variation 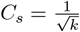.

At the start of each simulation, we set parameters dictating the levels of heterogeneity present in the population (*C*_*t*_, *C*_*s*_), the correlation between transmissibility and susceptibility (*ρ*), the average transmission rate 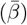, the recovery rate (*γ*), and the number of susceptible individuals and infected individuals at the start of the epidemic (*S*_0_ and *I*_0_). For our simulations, we set *C*_*t*_ = 0, 0.5, 1, or 3, *C*_*s*_ = 0, 0.5, 1, or 3, *ρ* = −1, − 0.5, 0, 0.5, or 1, 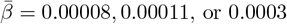, *γ* = 0.1, *S*_0_ = 999, and *I*_0_ = 1, giving a population size of *S*_0_ + *I*_0_ = 1, 000. We set 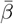 to correspond to *R*_0_ values of 0.8, 1.1, or 3, under the definition of *R*_0_ in Eq 6. We then randomly selected *I*_0_ individuals to start as infected, where each individual has the same probability of being selected (i.e., 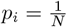 for all *i*).

We simulated the epidemic using a Gillespie algorithm. In this algorithm, we determine the time to the next event, which is when an infected individual either infects a susceptible individual or recovers. Infection occurs at rate 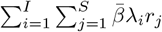, and recovery occurs at rate *γI*. If the next event is a new infection, we select a susceptible individual to become infected and move to the *I* class based on each individual’s relative risk such that the probability of selecting individual *i* is equal to 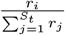. If the next event is a host recovery, we randomly select an infected individual to recover where each individual has an identical likelihood of being chosen. At each time point, we record the number of *S, I*, and *R* individuals as well as the effective reproductive number *R*_*e*_.

For each parameter combination of *C*_*t*_, *C*_*s*_, *ρ*, and *R*_0_, we ran 500 epidemic simulations. After simulating the epidemics, we calculated the following epidemic features for each simulation in which there was a major epidemic:

1. The probability of a major epidemic, defined as the fraction of simulations in which the final epidemic size is greater than a set threshold,
2. The peak size, defined as the maximum number of *I* individuals at any point in the epidemic,
3. The peak time, defined as the time at which the peak size occurs,
4. The final epidemic size, defined as the difference between the population size and the number of *S* individuals at the end of the epidemic *N* − *S*_final_,
5. The time to the *j*th infection, and
6. The effective reproductive number *R*_*e*_ defined at each time step *t* as in equation 8.

For a standard SIR model, the probability of a major epidemic can be calculated through a branching process approach, but the assumptions of a branching process model no longer hold when there are correlations between transmissibility and susceptibility and these traits vary continuously across individuals. Therefore, the threshold classifying an outbreak as a major epidemic was determined by plotting a histogram of the final epidemic sizes for the no correlation case and visually inspecting the histogram for the place where there was a clear division between epidemics in which few (minor) or many (major) individuals became infected (Southall et al., 2023). Because the level of heterogeneity in susceptibility impacts epidemic size (Gomes et al., 2022; Montalbán et al., 2022), we set a different threshold for different values of *C*_*s*_. Our threshold number of cases was 200 for *C*_*s*_ = 0.5, 100 for *C*_*s*_ = 1, and 50 for *C*_*s*_ = 3.

For each outbreak statistic, we determined the mean or median to compare the statistic across parameter combinations. Since the time to the *j*th infection is a time series with stochastic times, we first binned the output into 0.1 step size intervals to compare across simulations. To do so, we determined the status of this measure at each 0.1 step in time (i.e., at time 0, 0.1, 0.2,…) before calculating the median across the simulations at each time point over the epidemic.

### 4.2. Theoretical model results

In Fig 5, we show the trajectory of epidemics for *R*_0_ = 3 with different levels of heterogeneity in susceptibility, heterogeneity in transmission, and correlations between susceptibility and transmissibility, and we compare these trajectories to the case without heterogeneity and the case where susceptibility and transmissibility are uncorrelated. We found that correlations between transmissibility and susceptibility can greatly affect the probability of a major epidemic, peak size, peak time, final epidemic size, time to the *j*th infection, and *R*_*e*_. We also explored how different *R*_0_ may affect our results, using the Eq 6 definition of *R*_0_. Fig 6 shows epidemic trajectories for *R*_0_ = 0.8 and *R*_0_ = 1.1 with high levels of heterogeneity (*C*_*s*_ = *C*_*t*_ = 3). As expected, as *R*_0_ decreased, the probability of a major epidemic, peak size, and final epidemic size decreased. Also, the peak time and time to the *j*th infection tended to be later as *R*_0_ decreased.

**Figure 5:**
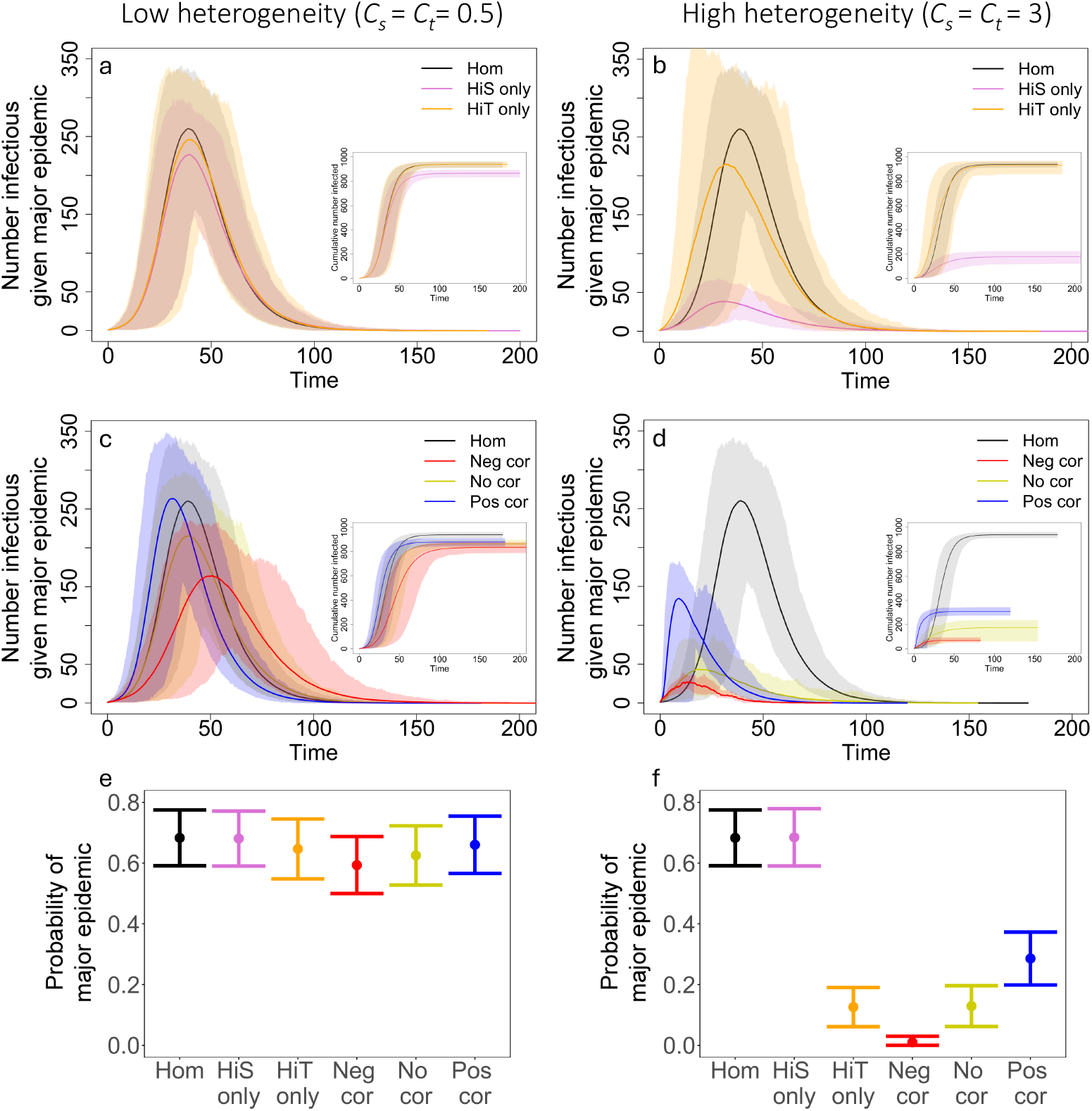
Correlations affect the disease dynamics in our model. The plots (a-d) show the average number of infected individuals over the course of an epidemic. Shaded regions represent 95% CIs determined from the major epidemics of 500 simulations for homogeneity (black), heterogeneity in susceptibility only (pink), heterogeneity in transmission only (orange), negative correlation (red, *ρ* = − 1), no correlation (yellow, *ρ* = 0), and positive correlation (blue, *ρ* = 1). (e,f) The average probability of a major epidemic for each case with 500 simulations and error bars of +/-2 standard deviations. (Insets) The cumulative number of individuals infected with 95% CIs from the major epidemics of 500 simulations. *C*_*s*_ = *C*_*t*_ = 0.5 in (a,c,e), *C*_*s*_ = *C*_*t*_ = 3 in (b,d,f), *N* = 1000, *I*_0_ = 1, and *R*_0_ = 3.

**Figure 6:**
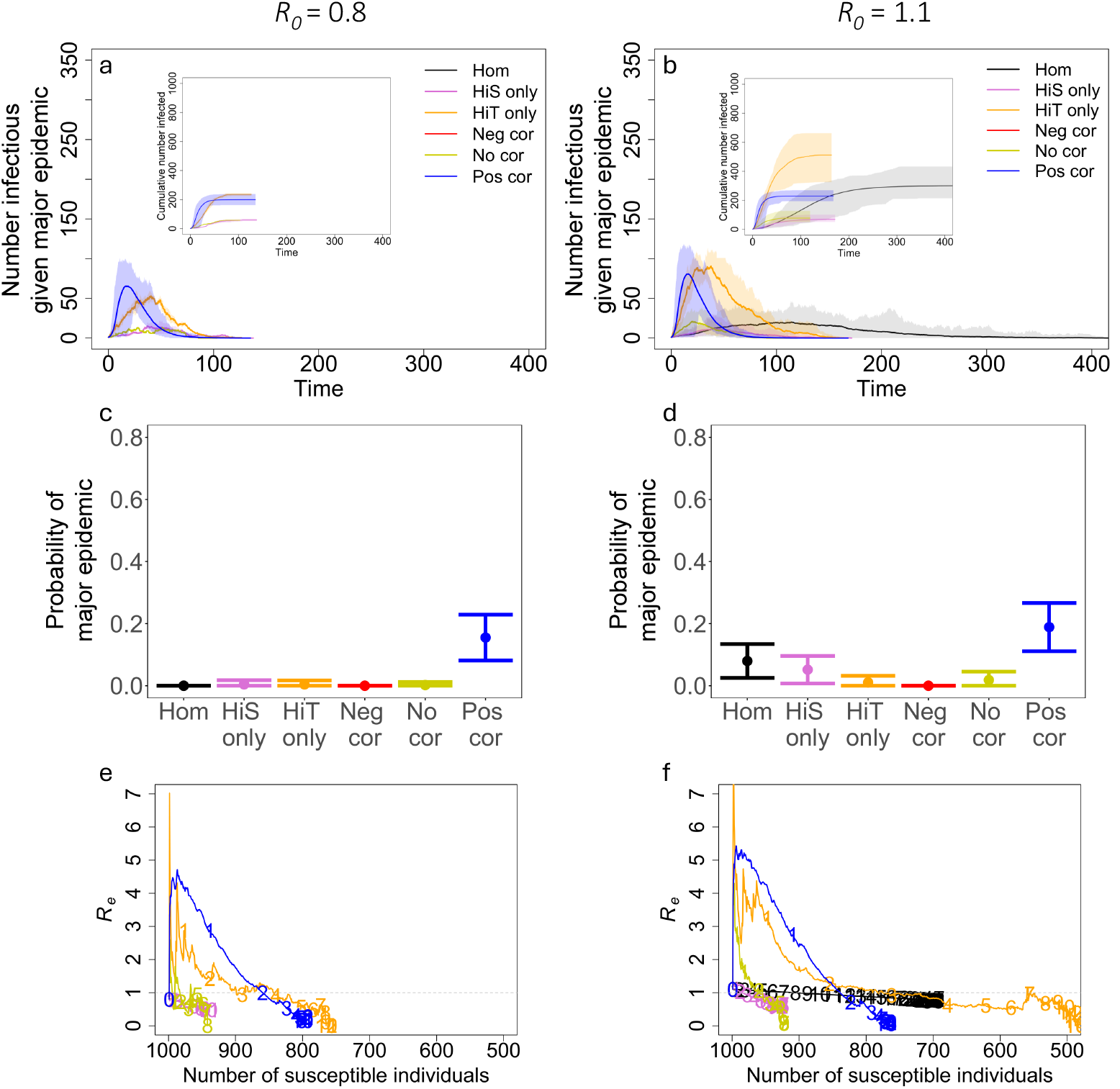
Positive correlations can lead to larger, more likely major epidemics when *R*_0_ is small. (a,b) The average number of infected individuals over the course of an epidemic. Shaded regions represent 95% CIs determined from the major epidemics of 500 simulations for homogeneity (black), heterogeneity in susceptibility only (pink), heterogeneity in transmission only (orange), negative correlation (red, *ρ* = −1), no correlation (yellow, *ρ* = 0), and positive correlation (blue, *ρ* = 1). (c,d) The average probability of a major epidemic for each case with 500 simulations and error bars of 2xSD. (e,f) *R*_*e*_ plotted against the number of susceptible individuals (*S*) averaged over the major epidemics from 500 simulations. The numbers on each trajectory represent time in the epidemic for every 10 units of time starting from the left (e.g., 1 is placed at time *t* = 10, 2 at *t* = 20, etc.). The dotted gray line shows *R*_*e*_ = 1. (Insets) The cumulative number of individuals infected with 95% CIs from the major epidemics of 500 simulations. *C*_*s*_ = *C*_*t*_ = 3, *R*_0_ = 0.8 in (a,c,e), *R*_0_ = 1.1 in (b,d,f), *N* = 1000, and *I*_0_ = 1.

Below we explore the effects of correlations between susceptibility and transmissibility on disease dynamics relative to the case with heterogeneities that are uncorrelated and the case where there is no heterogeneity (i.e., homogeneity). Generally, the direction of effects are the same at different values of *R*_0_, but see Supplement sections 4 and 5 for exceptions. We thus focus primarily on presenting results for the case where *R*_0_ = 3, but we specify when the results are different with smaller *R*_0_.

We first examined the effect of heterogeneity alone on disease dynamics. As expected based on previous studies, we found that with heterogeneity in susceptibility alone (*C*_*s*_ = 0.5, 1, or 3 and *C*_*t*_ = 0), epidemics had smaller peak sizes and smaller final epidemic sizes than epidemics without heterogeneity in susceptibility (i.e., homogeneity; Fig 5 and Supplement Figs S1 and S2). Also as expected, we found that heterogeneity in transmission alone (*C*_*t*_ = 0.5, 1, or 3 and *C*_*s*_ = 0) reduced the probability of a major epidemic (Fig 7). When epidemics did occur, they had the same peak and final epidemic sizes as the homogeneity case (Fig 5 and Supplement Figs S1 and S2).

**Figure 7:**
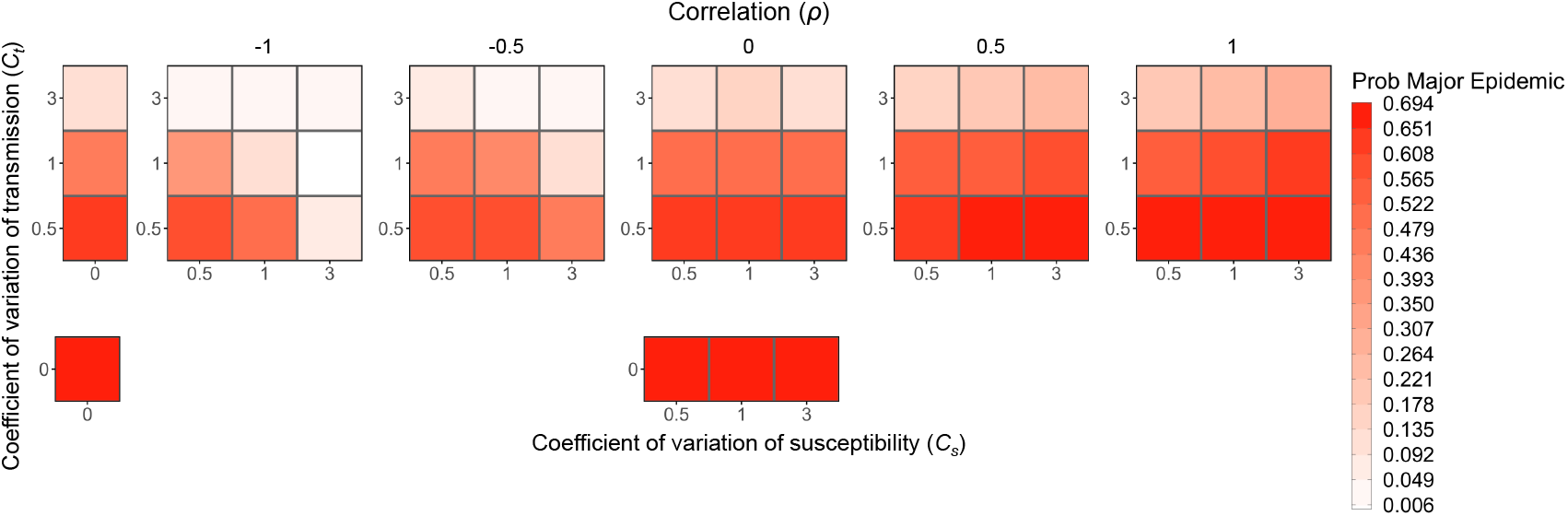
The probability of a major epidemic decreases as the level of heterogeneity in transmission increases, increases with positive correlation, and decreases with negative correlation. Each box is shaded to show the probability of a major epidemic, as defined in the text, averaged over the major epidemics from 500 simulations for various levels of heterogeneity in transmission (*C*_*t*_), heterogeneity in susceptibility (*C*_*s*_), and the correlation between transmissibility and susceptibility. While heterogeneity in transmission primarily determines the probability of a major epidemic, there are also effects from correlation and heterogeneity in susceptibility. Positive correlation results in a higher probability, negative correlation results in a lower probability, and increased levels of heterogeneity in susceptibility further decrease the chance of a major epidemic when there is a negative correlation. *N* = 1000, *I*_0_ = 1, and *R*_0_ = 3.

In a population heterogeneous with regard to both transmission and susceptibility, we found that when transmissibility and susceptibility are uncorrelated, the effect on disease dynamics is the same as combining the effects of each individual source of heterogeneity alone. In particular, when uncorrelated, the epidemics had a similar peak size and final epidemic size to those with heterogeneity in susceptibility alone (Fig 5 and respectively, Supplement Figs S1 and S2). Additionally, they had a similar probability of a major epidemic as with heterogeneity in transmission alone (Fig 7).

On the other hand, when there are correlations between transmissibility and susceptibility, these heterogeneities have interactive effects on disease dynamics. Since positive correlations indicate that the most susceptible individuals are also the most transmissible, when the first infected individual is transmissible enough to spread infection, epidemics readily take off and rapidly spread (Fig 5c and 5d). In this situation, however, epidemics also burn out quickly because the more susceptible individuals become infected early leaving only individuals that are lowly susceptible and lowly transmissible. In comparison to the uncorrelated case and the case without heterogeneity, a positive correlation results in a fast epidemic that quickly peaks and crashes. A positive correlation makes the overall probability of an epidemic more likely than in the uncorrelated case. Compared to the case where there is no heterogeneity, a positive correlation can make an epidemic less likely (Fig 5e and 5f) or more likely (Fig 6c and 6d) depending on the value of *R*_0_.

Since negative correlations indicate that the most susceptible individuals tend to be the least transmissible, and vice versa, this type of correlation between susceptibility and transmissibility causes epidemics to be hard to start and hard to sustain. This pattern arises because the individuals most likely to become infected are those least likely to cause new infections. When these epidemics do take off, in comparison to both the uncorrelated case and the case without heterogeneity, they are smaller, and they tend to either die out quickly or slowly meander through the population depending on the transmissibility of individuals infected early in the epidemic (Fig 5c and 5d).

As expected from previous literature (Lloyd-Smith et al., 2005) and our results with heterogeneity in transmission alone, the probability of a major epidemic is greatly influenced by the absolute magnitude of the heterogeneity in transmission (Fig 7). When holding correlations constant, increasing heterogeneity in transmission greatly decreases the chance of a major epidemic, but this effect disappears for subcritical *R*_0_ (Supplement Fig S13). Notably, correlations between susceptibility and transmissibility can partially offset the effect of heterogeneity in transmission, as demonstrated in Fig 7. In that figure and in Fig 5e and 5f, major epidemics occur more often with high heterogeneity in transmission and a positive correlation than with the same level of heterogeneity in transmission and no correlation (for example, the case where *C*_*t*_ = 3, *C*_*s*_ = 3, and *ρ* = 0 vs. *ρ* = 1). With decreased *R*_0_, the effects of correlations are even more pronounced. Positive correlations frequently lead to a greater chance of a major epidemic than in both the absence of correlation and the absence of heterogeneity with *R*_0_ = 1.1 (Supplement Fig S5). Moreover, with subcritical *R*_0_ = 0.8 and a positive correlation, major epidemics could still occur up to 26.2% of the time under the parameter sets explored (Supplement Fig S13). In the absence of correlations, there is virtually no impact on the probability of a major epidemic from the level of heterogeneity in susceptibility.

Also, as expected from previous literature (Aguas et al., 2020; Gomes et al., 2022; Montalbán et al., 2022) and our results with heterogeneity in susceptibility alone, in the absence of correlations, the peak size and final epidemic size are reduced as the level of heterogeneity in susceptibility is increased, while these features are largely independent of heterogeneity in transmission (Fig 5 and Supplement Figs S1 and S2). When susceptibility and transmissibility are correlated, however, changing the magnitude of heterogeneity in transmission can have large impacts on peak size and final epidemic size (Supplement Figs S1 and S2). Likewise, with positive correlations, it is typically the case that peak sizes are larger than the homogeneity case (Supplement Figs S1, S6, and S14), but the effect on the final epidemic size depends on the value of *R*_0_ (Supplement Figs S2, S7, and S15). These features arise with a positive correlation because the epidemic sweeps through the population quickly and crashes, typically resulting in a large peak but ultimately smaller epidemic (except when *R*_0_ is small). Negative correlations on the other hand lead to smaller peaks and final epidemic sizes than both the no correlation and homogeneous cases. This is because epidemics are hard to sustain with negative correlations since the individuals that are most likely to become infected tend to be least likely to transmit onward.

In contrast to the other epidemic features, which were driven primarily by levels of heterogeneity in either susceptibility or transmission, the peak time of an epidemic is strongly modified by the direction and magnitude of correlations between susceptibility and transmissibility rather than by the amount of heterogeneity itself (Fig 8). With positive correlations, epidemics take off quickly, so the peak tends to occur earlier in comparison to both the no correlation case and no heterogeneity case. With negative correlations, epidemics that take off tend to do so slowly, so the peak tends to occur later than in the no correlation and no heterogeneity cases. Epidemics that successfully take off in this negative correlation case, however, often start with a highly transmissible individual, so the peak can be earlier or later depending on the level of heterogeneity in transmission.

**Figure 8:**
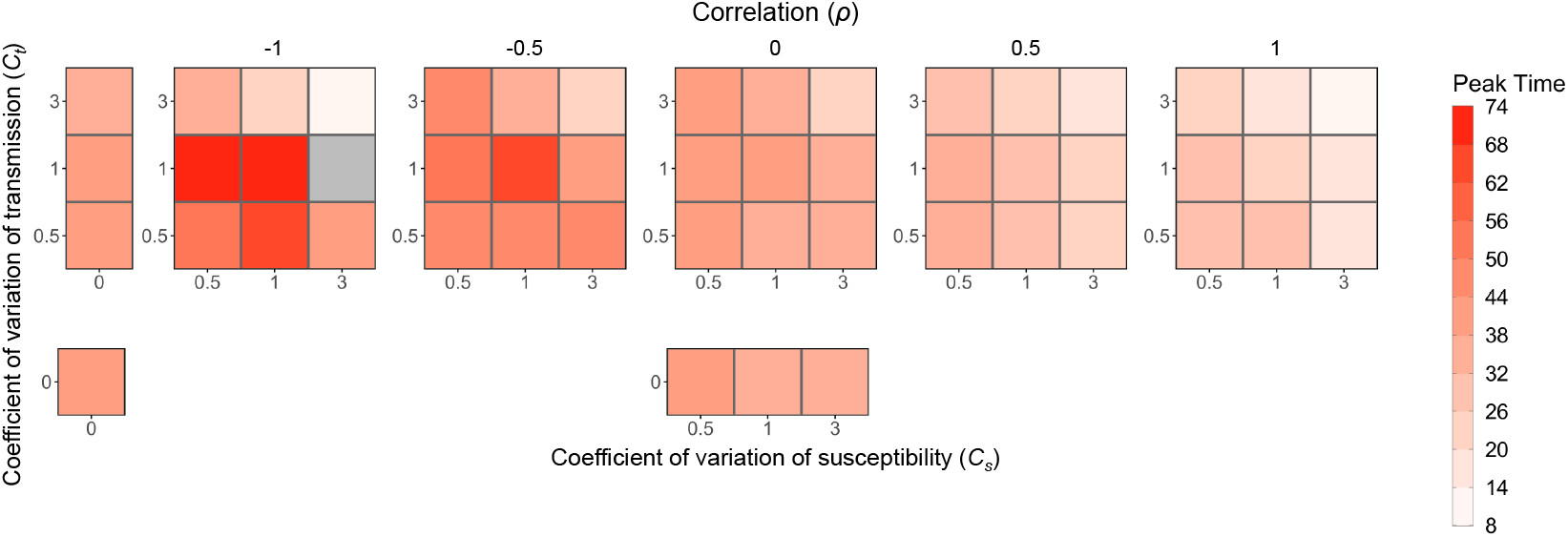
Peak time is earlier with positive correlation and later with negative correlation. Each box is shaded to show the peak time averaged over the major epidemics from 500 simulations for various levels of heterogeneity in transmission (*C*_*t*_), heterogeneity in susceptibility (*C*_*s*_), and the correlation between transmissibility and susceptibility. The number of simulations that were major epidemics, which can be determined by the probability of a major epidemic in Fig 7, was different for each parameter combination, so the average in each box is based on a different sample size. The gray box represents a parameter combination that resulted in no major epidemics as defined in the text. *N* = 1000, *I*_0_ = 1, and *R*_0_ = 3.

Another way to show the impact of correlation on the timing of an epidemic is to explore the time to the *j*th infection. Taking this approach reveals that the direction and magnitude of correlations between susceptibility and transmissibility can drive the timing of the *j*th infection, with patterns generally similar to those of peak time (Fig 9). We also simplified the data from Fig 9 to show only the time to the 50^th^ infection in order to compare across correlations and levels of heterogeneity, and we looked at the time for the epidemic to reach 50% of its final size (i.e., *j* = 0.5(*N* − *S*_final_)) to compare the speed of epidemics with different sizes and found generally similar results (Supplement Figs S3 and S4). Compared to the case with heterogeneity but no correlation, a positive correlation leads to the *j*th infection happening earlier and a negative correlation leads to the *j*th infection happening later. Notably, a positive correlation also leads to a faster epidemic than in the homogeneous case, but as can be seen in Fig 9, this changes for large values of *j* (i.e., where the *j*th infection occurs close to the end of the epidemic). As shown in Fig 9, a negative correlation results in a slower epidemic than other cases except when there is high heterogeneity in transmission (*C*_*t*_ = 3). In this case, the time to the *j*th infection for negative correlation is earlier than both the no correlation and homogeneity cases for small *j* but is later for large *j*. This is because the first infected individual must have high transmissibility for a major epidemic to occur, but the epidemic slows down over time. Fig 9 additionally shows that the trajectories for heterogeneity in transmission alone and no correlation tend to match those for homogeneity and heterogeneity in susceptibility alone respectively, which occurs because the difference in timing is primarily driven by heterogeneity in susceptibility or correlations between susceptibility and transmissibility.

**Figure 9:**
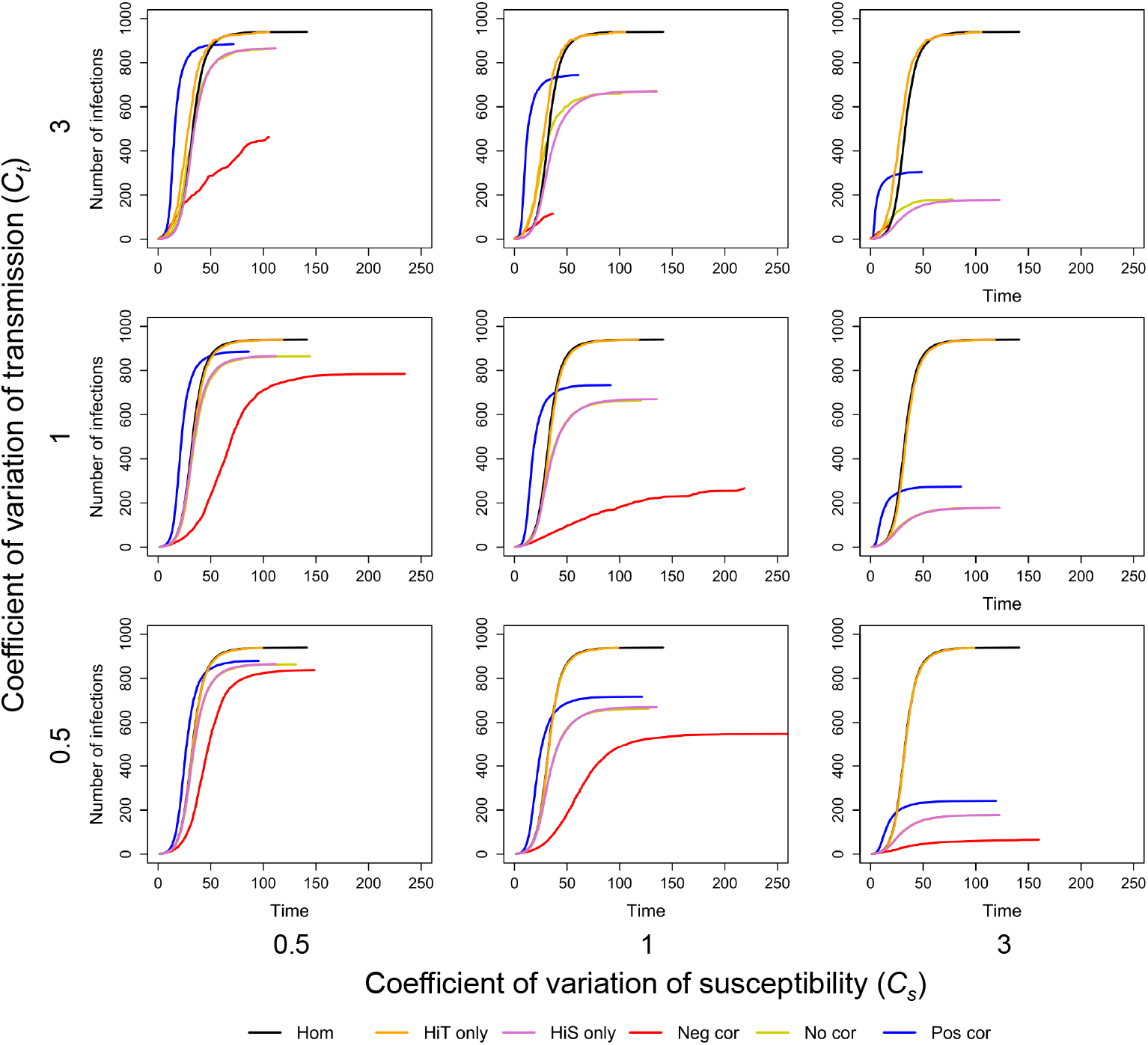
Time to the *j*th infection is earlier with positive correlation and later with negative correlation. The plots show the median time to the *j*th infection (where *j* is the value on the y-axis) from the major epidemics from 500 simulations for various levels of heterogeneity in transmission (*C*_*t*_), heterogeneity in susceptibility (*C*_*s*_), and the correlation between transmissibility and susceptibility. Each plot includes trajectories for the cases of homogeneity (black), heterogeneity in transmission alone (orange), heterogeneity in susceptibility alone (purple), perfect negative correlation (*ρ* = 1, red), no correlation (yellow), and perfect positive correlation (*ρ* = 1, blue). The number of simulations that were major epidemics, which can be determined by the probability of a major epidemic in Fig 7, was different for each parameter combination, so each line is based on a different sample size. There is no line for negative correlation with *C*_*t*_ = 1 and *C*_*s*_ = 3 because this parameter combination resulted in no major epidemics as defined in the text. *N* = 1000, *I*_0_ = 1, and *R*_0_ = 3.

In addition to these measures, we looked at the effect of correlations on the effective reproductive number *R*_*e*_ over the course of a major epidemic, by plotting *R*_*e*_ versus the number of susceptible individuals in the population *S* (Fig 10). This plot additionally shows how *R*_*e*_ changes over time. In the case with no heterogeneity, *R*_*e*_ is linearly related to the size of the susceptible population. When there is heterogeneity in transmission only, the average *R*_*e*_ follows the same trajectory with respect to *S* as in the homogeneous case although there is variation between outbreak simulations. When there is heterogeneity in susceptibility only, *R*_*e*_ declines faster with respect to *S* than in the case with no heterogeneity, but it follows the same trajectory as the no correlation case. This is because the presence of heterogeneity in susceptibility creates an infection selection process whereby highly susceptible individuals are infected early in the epidemic, resulting in an increasingly less susceptible population and therefore smaller *R*_*e*_ over time. The dynamics are fundamentally altered by positive and negative correlations between susceptibility and transmissibility. In the presence of a positive correlation, *R*_*e*_ shoots up at the start of an epidemic before rapidly declining faster than linearly against *S*. In addition, the trajectory plays out faster in time than in the other cases examined. This is because the epidemic quickly sweeps through the most susceptible and transmissible individuals before rapidly dying out. In the presence of a negative correlation, *R*_*e*_ immediately drops then rapidly declines as a function of *S*. The exception to this is that when there is high heterogeneity in transmission (*C*_*t*_ = 3), *R*_*e*_ first shoots up before dropping. This happens because major epidemics only occur in this case when the first infected individual has high transmissibility. The rate of decline of *R*_*e*_ as a function of time, however, is slower with negative correlations than in the other cases after the initial precipitous drop. These patterns arise because individuals that are more susceptible do not contribute as much to onward transmission and vice versa, resulting in a slower epidemic. With *C*_*t*_ = 3, however, major epidemics may be faster for negative correlations because the individual infected at the start must have high transmissibility for the epidemic to take off.

**Figure 10:**
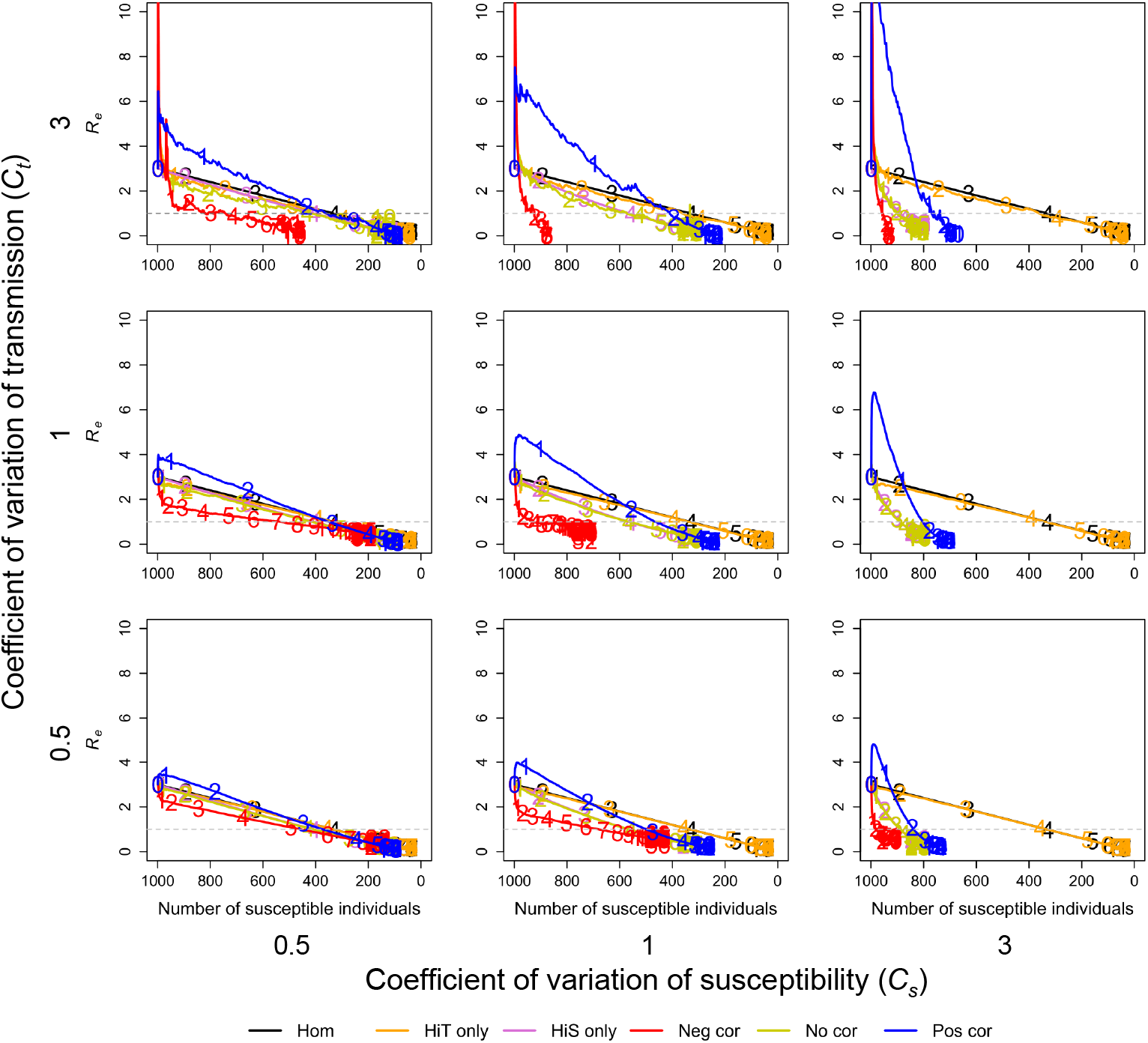
The effective reproductive number *R*_*e*_ depends on both the levels of heterogeneity and the correlation. The plots show *R*_*e*_ plotted against the number of susceptible individuals (*S*) averaged over the major epidemics from 500 simulations for various levels of heterogeneity in transmission (*C*_*t*_), heterogeneity in susceptibility (*C*_*s*_), and the correlation between transmissibility and susceptibility. Each plot includes trajectories for the cases of homogeneity (black), heterogeneity in transmission alone (orange), heterogeneity in susceptibility alone (purple), perfect negative correlation (red, *ρ* −= 1), no correlation (yellow, *ρ* = 0), and perfect positive correlation (blue, *ρ* = 1). The numbers on each trajectory represent time in the epidemic for every 10 units of time starting from the left (e.g., 1 is placed at time *t* = 10, 2 at *t* = 20, etc.). The dotted gray lines show *R*_*e*_ = 1. The number of simulations that were major epidemics, which can be determined by the probability of a major epidemic in Fig 7, was different for each parameter combination, so each line is based on a different sample size. There is no line for negative correlation with *C*_*t*_ = 1 and *C*_*s*_ = 3 because this parameter combination resulted in no major epidemics as defined in the text. *N* = 1000, *I*_0_ = 1, and *R*_0_ = 3.

## 5. Application to mpox

Thus far, we have focused on exploring the effects of correlations between susceptibility and transmissibility on disease dynamics in a theoretical model. In 2022, there was a global outbreak of mpox, with a notable explosive outbreak occurring in New York City (Kaftan et al., 2024; Pekar et al., 2025). Since transmission in this outbreak was largely attributed to behaviors such as sexual encounter rates, it is reasonable to expect risk and transmission potential to vary between individuals, generating heterogeneity in susceptibility and transmission. Moreover, it is reasonable to think that the most at risk individuals would also be the most transmissible once infected. Mpox therefore represents a system in which there might be substantial heterogeneity and strong positive correlations between susceptibility and transmissibility. It therefore serves as a useful test case of the theory.

### 5.1. Mpox methods

To explore the impacts of correlations between susceptibilty and transmissibility on disease dynamics in real systems, we expanded our simple SIR model to more accurately represent the dynamics of mpox. The key changes necessary to make this possible were to introduce an exposed class of hosts (since mpox dynamics can be impacted by an incubation period between exposure and infectiousness) and to have new hosts enter and leave the population over time for non-disease related reasons. We thus expanded our SIR model into an SEIR model:

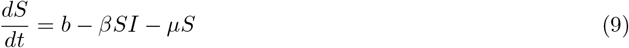

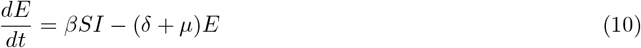

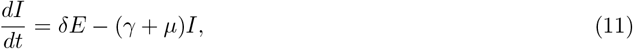

where all parameters are as described in the standard SIR model with the following new additions: *E* is the number of exposed individuals, *b* is the rate at which individuals enter the at-risk population, and *µ* is the rate at which individuals leave the at-risk population. Note that this model is not intended to track all individuals in New York City but rather only the individuals considered to be at-risk for mpox. Even within this group of at-risk individuals, individuals may differ in their transmissibility and susceptibility due to differences in behavior. We thus included heterogeneity in and correlation between transmissibility and susceptibility as described above in our theoretical model setup.

We compared this model output to the 2022 mpox outbreak in New York City, which was the epicenter of the US mpox outbreak with its first mpox case tested on May 19, 2022 and confirmed positive on May 21, 2022 (NYC DOHMH, a). To do so, we used parameter estimates generated from previously published data on this mpox outbreak and others (Blumberg and Lloyd-Smith, 2013; Branda et al., 2023; Gao et al., 2023; Brand et al., 2023; Kaftan et al., 2024). We then compared the model predictions to mpox case count datasets publicly available on GitHub (NYC DOHMH, b) and listed on the New York City mpox information website (NYC DOHMH, a) from May 19, 2022 to March 8, 2025. Cases were counted on their date of specimen collection rather than their date of reporting. As mpox is spread mainly through close contact, we set a perfect positive correlation *ρ* = 1 between susceptibility and transmissibility (Laurenson-Schafer et al., 2023), which would occur if the individuals who had the most contacts prior to infection also had the most contacts after infection. We additionally analyzed the effect that correlation itself is having in this system by generating alternative models in which we set *ρ* = 0 and *ρ* = −1. Assuming a high level of heterogeneity between hosts, we set *C*_*t*_ = *C*_*s*_ = 3, which is roughly consistent with prior estimates of mpox heterogeneity (Blumberg and Lloyd-Smith, 2013). We set *δ* = 0.13 and *γ* = 0.071 based on values used in a forecasting model of mpox in New York City (Kaftan et al., 2024). Following this same model, we assumed the at-risk host population size to be 70, 180 with 19 infected individuals on May 2, 2022 (Kaftan et al., 2024). Based on prior estimates of *R*_0_ for mpox (Branda et al., 2023; Gao et al., 2023; Brand et al., 2023; Kaftan et al., 2024), we set *R*_0_ to be 0.52 with positive correlation using the *R*_0_ definition from Eq 6, which corresponds to an *R*_0_ of 5.2 following Eq 7. In practice, this was achieved by setting 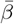 to 5.4 × 10^7^. We then compared the effects of correlation in this system by keeping either the initial mean transmissibility and mean susceptibility constant (i.e., *R*_0_ defined as in Eq 6) or by keeping the mean product of transmissibility and susceptibility constant (i.e., *R*_0_ defined as in Eq 7). We set *b* = 70 to account for individuals entering the at-risk population due to factors such as a change in behavior or migration into New York City. To balance total population size in the at-risk population, we set the rate of migration out at *µ* = 0.000997 to maintain a constant population size. This is roughly equivalent to the average individual remaining in the at-risk population for 2.75 years.

We simulated the mpox epidemic following a similar Gillespie framework as described above except that there were now more possible events. The possible events in this mpox model are that a susceptible individual can enter the population, an infected individual can infect a susceptible individual, an exposed individual can become infectious, an infected individual can recover, or an individual of any type can leave the population. Infection and recovery occurred at the same rate and in the same way as in the theoretical model above, except that infected individuals now moved from the *S* to the *E* class. Across the whole population, individuals entered the population at rate *b*, exposed individuals become infectious at rate *δE*, and individuals left the population at rate *µ*(*S* + *E* + *I* + *R*). When the next event was an individual entering the population, we added a new susceptible individual and assigned this individual a relative force of infection *λ*_*i*_ and a relative risk of being infected *r*_*i*_ as detailed above. When the next event was an exposed individual becoming infectious, we randomly selected an exposed individual to move to the *I* class where each individual had an identical likelihood of being chosen. When the next event was an individual leaving the population, we randomly selected an individual to remove where all individuals in the population had identical likelihoods of being chosen. We ran 500 simulations for each level of correlation (*ρ* = −1, 0, 1). After simulating the mpox epidemic, we applied a case detection rate of 5%, which is consistent with prior estimates of the case detection rate (Kaftan et al., 2024). We then calculated the mean and 95% CI at each time point.

### 5.2. Mpox results

The 2022 mpox outbreak in New York City was remarkable for how quickly it started, how quickly case counts grew, and how abruptly it ended despite there presumably still being a large pool of susceptible individuals remaining. Figure 11 shows daily case counts of mpox in New York City and the trajectory of mpox as described by our SEIR model with positive, zero, or negative correlations between transmissibility and susceptibility. Notably, these features of the outbreak are generally consistent with our model that assumes positive correlation between transmissibility and susceptibility. Our model with positive correlations additionally does a reasonably good job of capturing the outbreak peak, the overall case counts, the long term persistence of mpox at low levels after the peak, and the small resurgence in case counts that occurs in 2024. However, when correlations are absent or negative, outbreaks simply do not occur at realistic values of *R*_0_. In the supplement, we further show that even when using unrealistic values of *R*_0_, this combination of features is not achieved in models that have no correlation or negative correlation between susceptibility and transmissibility (Supplement Fig S21). Overall, these results demonstrate the impact correlations can have on disease dynamics. Although our model with positive correlation does not perfectly capture the decline in cases between August and November of 2022, this slight discrepancy can be readily explained by vaccination and other public health measures that were imposed during the height of the outbreak, with the first mpox vaccine doses becoming available in New York City on July 6, 2022 (NYC DOHMH, 2022).

**Figure 11:**
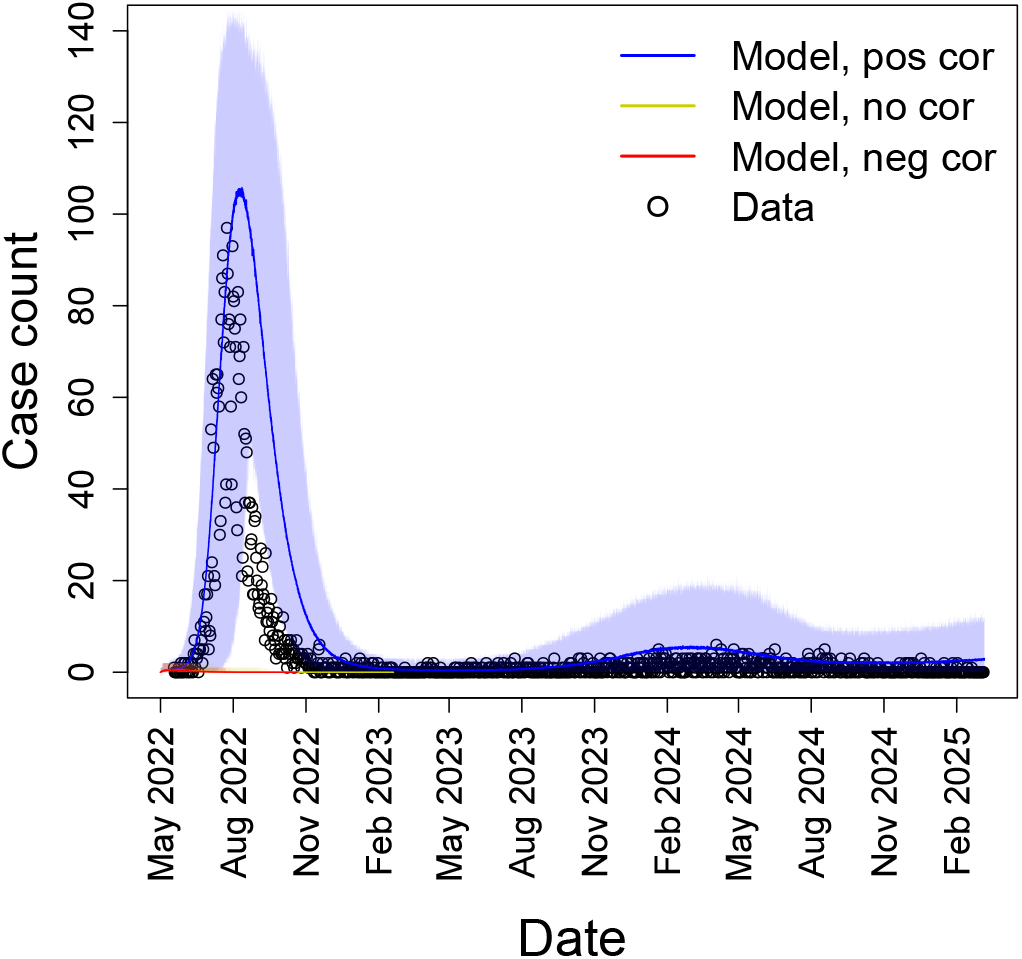
Mpox dynamics in New York City are generally consistent with positive correlations between transmissibility and susceptibility. Daily case counts of mpox in New York City (black circles) from May 19, 2022 to March 8, 2025 and the average number of infectious individuals from our SEIR model starting May 2, 2022 for positive correlation (blue, *ρ* = 1), no correlation (yellow, *ρ* = 0), and negative correlation (red, *ρ* = −1). Shaded regions represent the 95% CIs of 500 simulations. Note that there was a slightly faster than expected decline in cases compared to our positive correlation model in the latter half of the 2022 epidemic, which may be attributable to vaccination and other public health measures. *C*_*s*_ = *C*_*t*_ = 3, *R*_0_ = 0.52 (Eq 6), *N* = 70180, and *E*_0_ = 19.

## 6. Discussion

An important question in epidemiology is why epidemics follow different trajectories. It is well known that heterogeneities in transmission and susceptibility can individually impact disease dynamics in known ways (Dwyer et al., 1997; Lloyd-Smith et al., 2005; Gomes et al., 2014; Langwig et al., 2017; Gomes et al., 2022). Here we have shown that when both heterogeneities are present simultaneously, correlations between transmissibility and susceptibility can drastically alter disease dynamics. We showed this by first conducting a systematic review to identify what was previously known and what knowledge gaps remained, second by creating a stochastic, individual-based SIR model to generate results to fill in the knowledge gaps, and third, by applying an SEIR version of our theoretical model to a 2022 outbreak of mpox. Our literature review found that the effects of correlations between transmissibility and susceptibility were understudied, particularly with regard to how the dynamics of models with these correlations differed from those of models with heterogeneities but no correlations. Using our model, we found that if the drivers of heterogeneity in transmission and susceptibility are independent, then the effects on disease dynamics are essentially the same as each heterogeneity source alone (i.e., heterogeneity in transmission reduces the probability of a major epidemic, and heterogeneity in susceptibility reduces the peak and final epidemic sizes). In contrast, the impacts on disease dynamics are more complicated when susceptibility and transmissibility are correlated. Positive correlations result in major epidemics that are larger, faster, and more likely in comparison to the case with no correlation (Fig 12). In comparison to the case with no heterogeneity, epidemics are faster with positive correlations. When *R*_0_ is a bigger value, positive correlations lead to smaller and less likely major epidemics than the dynamics with homogeneity (Fig 5), but when *R*_0_ is close to or less than 1, positive correlations can lead to larger and more likely major epidemics (Fig 6). Negative correlations result in major epidemics that are less likely and smaller than both the no correlation and homogeneous cases, but these epidemics can be faster or slower than both cases depending on the level of heterogeneity in transmission (Fig 9). We showed these effects of correlations in a real system by applying our model to mpox in New York City (Fig 11). We found that the 2022 epidemic, as well as mpox dynamics through 2025, could be reasonably well explained by a model that includes a positive correlation between susceptibility and transmissibility but not by similar models that lack this positive correlation.

**Figure 12:**
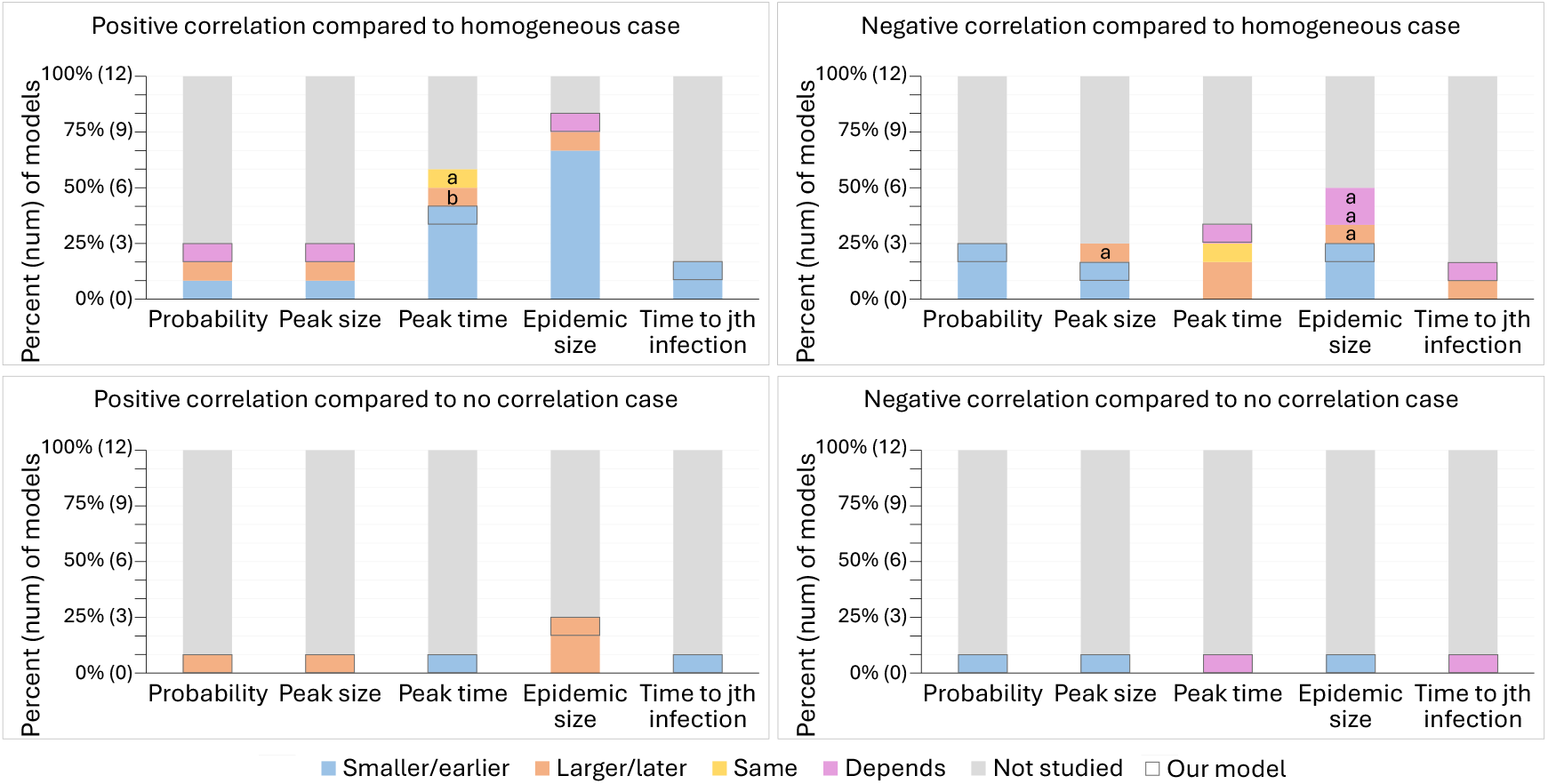
Results from our model fill in knowledge gaps about the effects of correlation from the systematic review. These plots show the effects of positive and negative correlations between transmissibility and susceptibility on the probability of a major epidemic, peak size, peak time, final epidemic size, and time to the *j*th infection in comparison to disease dynamics with homogeneity or no correlation according to 11 models from the 9 studies included in the systematic review and our model. The effect of correlation is classified for each model and measure as resulting in an attribute that is smaller/earlier (blue), larger/later (orange), the same (yellow), any of these results depending on the levels of heterogeneity or value of *R*_0_ (purple), or not studied (gray) in comparison to the homogeneous or no correlation case. The results from our model have a black outline. For positive correlation compared to the homogeneous case, we found that the effect of correlation on the probability of a major epidemic and the final epidemic size depends on *R*_0_ (defined by Eq 6), and the effect of correlation on the peak size depends on *R*_0_ and the level of heterogeneity. When *R*_0_ is close to or less than 1 (*R*_0_ = 0.8 or 1.1), positive correlation results in a larger probability, peak, and final epidemic size than with homogeneity, whereas when *R*_0_ is increased (*R*_0_ = 3), positive correlations results in a less likely and smaller epidemic than with homogeneity. With *R*_0_ = 3, the peak size can still be larger for positive correlations than homogeneity when there is high heterogeneity in transmission and low heterogeneity in susceptibility. Also, note that the time to the *j*th infection is earlier for positive correlations than homogeneity under the condition that *j* is not large (i.e., *j* is not close to the final epidemic size). For negative correlation, we found that the effect of correlation on both the peak time and the time to the *j*th infection depends on the level of heterogeneity in transmission. High heterogeneity leads to an earlier peak and *j*th infection compared to the homogeneous and no correlation cases while low heterogeneity leads to these attributes being later. The letters ’a’ and ’b’ denote results from the systematic review models that are inconsistent with our results where ’a’ means the model associated with that result defined *R*_0_ according to Eq 7 and ’b’ means the model kept a constant growth rate for the first month.

From our systematic literature review, we have identified that few published studies explored the impact of correlation between transmissibility and susceptibility on disease dynamics (Cardell and Kanouse, 1989; Becker and Marschner, 1990; Daley et al., 2000; Andreasen, 2011; Clancy and Pearce, 2013; Hickson and Roberts, 2014; Tkachenko et al., 2021; Kawagoe et al., 2021; Allard et al., 2023). The studies that do exist tend to have compared models with heterogeneity and correlations to models without heterogeneity, meaning that they were largely unable to assess the impact of the correlation itself on disease dynamics. Moreover, the studies that have been published used different model structures, parameter values, and assumptions, resulting in disagreements on the effects of correlations not just in magnitude but also in direction. We found that the majority of models agreed that positive correlations resulted in smaller final epidemic sizes and earlier peaks in comparison to cases without heterogeneity (Cardell and Kanouse, 1989; Daley et al., 2000; Andreasen, 2011; Clancy and Pearce, 2013; Hickson and Roberts, 2014; Tkachenko et al., 2021; Kawagoe et al., 2021). In two models cases, however, this did not occur (Cardell and Kanouse, 1989; Hickson and Roberts, 2014). For one of the models, Cardell and Kanouse (1989) kept the growth rate constant for the first month of the epidemic to compare between cases, resulting in a later peak as individuals not infected during the first month are more likely to be both less susceptible and less transmissible. In the other case, Hickson and Roberts (2014) kept the mean product of transmissibility and susceptibility constant across cases and thus found positive correlations resulted in a smaller epidemic size but a similar peak time compared to homogeneity. Our literature review also found disagreement between the effects of negative correlations on disease dynamics. For instance, in comparison to models without heterogeneity, some models found that negative correlations could result in a smaller (Daley et al., 2000; Andreasen, 2011; Clancy and Pearce, 2013; Hickson and Roberts, 2014), larger (Andreasen, 2011; Clancy and Pearce, 2013; Hickson and Roberts, 2014), or similar (Andreasen, 2011; Clancy and Pearce, 2013) final epidemic size. This may be because of the respective authors’ choices of distributions used to model susceptibility and transmissibility or the metrics kept constant to make comparisons across cases. Lastly, our literature review revealed that the vast majority of prior studies only compared models with correlations to models without heterogeneity, and thus neglected the comparison to models with heterogeneity but no correlation between transmissibility and susceptibility. The two that compared the case with correlations to the case without correlations restricted their analysis to only final epidemic size and a positive correlation, meaning that there were substantial knowledge gaps in understanding how the correlation between susceptibility and transmissibility impacted disease dynamics (Fig 4).

There has recently been renewed interest in understanding how heterogeneity in and interactions between traits related to transmissibility and susceptibility affect disease dynamics. Beagle et al. (2022) investigated how correlations between traits related to transmissibility and superspreading affect peak epidemic size. They found that, in comparison to the case without correlations, positive correlations between contact rate and infectiousness result in larger peak sizes, whereas negative correlations result in smaller peak sizes. We believe that this result arose in their analysis because, despite their focus on transmissibility, contact rate also implicitly impacts susceptibility. Their results thus further support our finding that correlations between susceptibility and transmissibility impact peak size (increase with positive correlations and decrease with negative correlations).

In a recent preprint, Harris et al. (2024) also investigated the effects of correlations between transmissibility and susceptibility using truncated Gaussian distributions with correlations ranging from −0.6 to 0.6. Largely in agreement with our results, they found that in comparison to the case with no correlation between susceptibility and transmissibility, positive correlations lead to faster, stronger, more likely epidemics, whereas negative correlations lead to slower, weaker, less likely epidemics. Our analysis differs from theirs in that we used gamma distributions for our heterogeneity, we analyzed the impact of correlations ranging from −1 to 1, and we investigated a wider range of values for the level of heterogeneity. In addition, we explored the effect of altering *R*_0_, which demonstrated that positive correlations can lead to larger major epidemics compared to the homogeneous case even in cases where no outbreak would occur in the absence of heterogeneity.

*R*_0_, or the basic reproductive number, is a benchmark parameter used to evaluate the threat posed by an emerging infectious disease. While this number can be, and has been, valuable for predicting and managing epidemics (Ferguson et al., 2001; Lipsitch et al., 2003; Hens et al., 2015), its interpretation can change in the presence of heterogeneity. The probability of a major epidemic can be calculated based on *R*_0_, but this probability must be adjusted if populations have heterogeneity in transmission (Lloyd-Smith et al., 2005). Likewise, the final epidemic size and threshold for herd immunity can be calculated based on *R*_0_, but these values must be adjusted to account for heterogeneity in susceptibility (Ball, 1985; Montalbán et al., 2022). We have shown that when susceptibility and transmissibility are correlated, further adjustments are needed. These issues are exacerbated by small details that impact the calculation of *R*_0_. As we discussed above, *R*_0_ should be defined as the product of mean transmissibility and mean susceptibility when individuals are equally likely to be the first infected (Eq 6), but *R*_0_ should be alternatively defined by the mean product of transmissibility and susceptibility when the likelihood of being the first infected is exactly proportional to an individual’s susceptibility (Eq 7). These values need not be similar and holding one versus the other constant can lead to fundamentally different conclusions regarding the effects of correlations between susceptibility and transmissibility on disease dynamics. Four of the 11 models in our systematic review made comparisons across disease dynamics by keeping the initial mean transmissibility and mean susceptibility constant (i.e., *R*_0_ defined as in Eq 6). Five of the 11 models made comparisons by keeping the initial mean product of transmissibility and susceptibility constant (i.e., *R*_0_ defined as in Eq 7). This reflects the ambiguity present in defining and interpreting *R*_0_. For our analysis, we used the first definition of *R*_0_ (Eq 6), electing to keep the mean force of infection and mean risk of infection constant between different levels of correlations and heterogeneity. One way this played out is that the effective reproductive number *R*_*e*_ immediately differed between the positive, negative and uncorrelated cases. It also led to the counterintuitive results that major epidemics could still occur, and with relatively high probability, when *R*_0_ *<* 1 in the case of positive correlations because *R*_*e*_ typically became greater than 1 after the first transmission event. Hence, *R*_0_, as defined by Eq 6, is a poor measure of outbreak potential when there are correlations between transmissibility and susceptibility.

We propose that incorporating heterogeneity and correlations into disease models may improve predictive capability, particularly in cases where there are known or likely correlations between transmissibility and susceptibility. For instance, variation in the number of contacts that individuals have is a frequently assumed mechanism for generating heterogeneity in transmission (Lloyd-Smith et al., 2005). In such cases, there is an implicit positive correlation between susceptibility and transmissibility since contact rate impacts both exposure and onward transmission. However, other sources of heterogeneity can be difficult to estimate, especially early in an epidemic. This is slowly changing, since recent increased interest in real-time disease forecasting has resulted in the development of novel methods to estimate heterogeneity in susceptibility (Gomes et al., 2022; Anderson et al., 2023; Tuschhoff and Kennedy, 2024). We propose that it will likewise be important to develop methods to measure correlations between susceptibility and transmissibility.

Based on our finding that positive correlations lead to larger, faster, more likely major epidemics, we suggest that increased surveillance for disease outbreaks may be warranted in places or subpopulations where positive correlations are likely. Likewise, public health interventions can be targeted towards settings where positive correlations are likely, and ideally, interventions could be aimed at attempting to induce negative correlations. An example of this would be targeting limited vaccines towards individuals with many contacts. This strategy has been proposed previously for the purpose of reducing superspreading events without consideration for correlation (Vidondo et al., 2012; Manzo and van de Rijt, 2020; Saunders and Schwartz, 2021). If such correlations were present, taking these correlations into account would increase the support in favor of utilizing this approach.

One consideration of our study is that we investigated the effect of correlation between two variables (the force of infection and risk of infection) on disease dynamics. In reality, there may be many interacting variables with varying degrees and directions of correlation. For example, there may be correlations between factors associated with infection duration, such as recovery rate or virulence, and those important for disease transmission, such as the force of infection and risk of infection that we investigated. The impact of this added complexity may affect disease dynamics differently than what we found (Beagle et al., 2022) and should be explored further.

Overall, the correlation between transmissibility and susceptibility is an important consideration for epidemiological models since all of our epidemic measures were affected by correlation to some extent. Although the probability of a major epidemic, peak size, and final epidemic size were greatly affected by the levels of heterogeneity, positive correlations generally counteracted the effects of heterogeneity and negative correlation generally enhanced them. The impact of a positive correlation is especially important for populations with small or subcritical *R*_0_ where it is still possible to have sizable major epidemics even when no such epidemic would occur without a correlation or in the absence of heterogeneity. The timing and speed of an epidemic (i.e., the peak time and time to the *j*th infection) were driven largely by correlations, with a positive correlation leading to a faster epidemic and a negative correlation leading to a slower one. The levels of heterogeneity also impacted timing, particularly early in the epidemic, where increased heterogeneity in transmission led to a more explosive epidemic, and increased heterogeneity in susceptibility also led to a faster epidemic as infection swept through the more susceptible individuals. As all of these measures of disease dynamics are wrapped up with *R*_*e*_, both correlation and the levels of heterogeneity strongly influenced *R*_*e*_.

As we demonstrated, the 2022 mpox epidemic may provide a real-world example of the effect of correlations. In that outbreak, the number of mpox cases rapidly increased and then rapidly declined after only a small fraction of the overall population was infected (Murayama et al., 2024). Our model of mpox in New York City does a reasonably good job of capturing the initial growth of cases, the peak size and time, and the eventual long term persistence of mpox at low levels when we include positive correlations between transmissibility and susceptibility. While other factors may have contributed, such as information campaigns and targeted interventions (Delaney et al., 2022), the rapid rise and subsequent decline of mpox is consistent with the dynamics caused by positive correlations between transmissibility and susceptibility. It still remains an open question to determine the degree to which the control of mpox was due to public health measures versus the epidemic running its course (Murayama et al., 2024; Pekar et al., 2025), but our modeling results suggest it is at least possible that the epidemic had nearly run its course prior to the implementation of a public health response.

## Data Availability

All code used to produce and analyze these results is publicly available on a GitHub repository at https://github.com/bmtuschhoff/correlations.git.

## Acknowledgements

We thank Ephraim Hanks for feedback on our model setup. DAK and BMT were supported by Institute of General Medical Sciences, National Institutes of Health (R01GM140459) and the UK Biotechnology and Biological Sciences Research Council as part of the joint NSF-NIH-USDA Ecology and Evolution of Infectious Diseases program. DAK was also supported by National Science Foundation grant DEB-2211322. The funders had no role in study design, data collection and analysis, decision to publish, or preparation of this article.

## Notes

### Competing Interest Statement

The authors have declared no competing interest.

### Summary of Updates

Section 2 revised to introduce SIR models before introducing R0; Table 1 added, which defines all model parameters; Four figures moved from the main text to the supplement (Figs S1, S2, S3, S13); Section 5 added to apply an SEIR version of our theoretical model to mpox disease dynamics in New York City; Supplement updated

