## Supplement for "Heterogeneity in and correlation between host transmissibility and susceptibility can greatly impact epidemic dynamics"

---

### 1. Systematic review

Table S1: The model structure, assumptions, and results for positive correlation associated with each of the 11 models included in the systematic review and our model. For model structure, the model type is either an SIR-type (SIR, SI, etc.), a branching process, an age of infection model, or a directed network. Models were set up as deterministic, stochastic, or both to explore different results. The distributions used to model the values for susceptibility and transmissibility were either discrete or continuous and the type of distribution explored in each study was a gamma distribution, an unnamed distribution where authors set their own values, a gamma distribution plus another distribution, or solely another type of distribution like a lognormal or uniform distribution. For assumptions, models compare the effects of correlation to the uncorrelated and homogeneous cases by keeping the mean susceptibility and mean transmissibility constant ("Mean $r$ and $\lambda$"), keeping the mean product of susceptibility and transmissibility constant ("Mean $r * \lambda$"), or by keeping the growth rate for the first month of the epidemic or the median product of susceptibility and transmissibility constant ("Other"). Most models assume a well-mixed population and changing susceptible population size over time. The effect of correlation is classified for each model and measure as resulting in an attribute that is smaller/earlier (blue), larger/later (orange), the same (yellow), any of these results depending on the levels of heterogeneity or value of $R_0$ (purple), or not studied (gray) in comparison to the homogeneous or no correlation case. Boxes with lighter shading indicate that the corresponding model did not explicitly conclude that effect, but it could be determined from the results presented. For positive correlation compared to the homogeneous case, we found that the effect of correlation on the probability of a major epidemic and the final epidemic size depends on $R_0$, and the effect of correlation on the peak size depends on $R_0$ and the level of heterogeneity. When $R_0$ is close to or less than 1 ($R_0 = 0.8$ or 1.1), positive correlation results in a larger probability, peak, and final epidemic size than with homogeneity, whereas when $R_0$ is increased ($R_0 = 3$), positive correlations results in a less likely and smaller epidemic than with homogeneity. With $R_0 = 3$, the peak size can still be larger for positive correlations than homogeneity when there is high heterogeneity in transmission and low heterogeneity in susceptibility.

| Paper Information | | Model Structure | | | | Assumptions | | | | Results for positive correlation | | | | | | | | | |
|---|---|---|---|---|---|---|---|---|---|---|---|---|---|---|---|---|---|---|---|
| | | | | | | | | | | Probability of major epidemic | | Peak size | | Peak time | | Final epidemic size | | Time to jth infection | |
| First author | Year | Model type | Deterministic? | Discrete values? | Distribution | Constant comparison | Well-mixed | Changing S | Type of correlation studied | Compare to hom | Compare to no cor | Compare to hom | Compare to no cor | Compare to hom | Compare to no cor | Compare to hom | Compare to no cor | Compare to hom | Compare to no cor |
| NS Cardell | 1989 | SIR-type | Deterministic | Continuous | Gamma | Mean r and λ | Yes | Yes | Positive | x | x | x | x | Earlier | x | Smaller | x | x | x |
| NS Cardell | 1989 | SIR-type | Deterministic | Continuous | Gamma | Other (growth rate for first month) | Yes | Yes | Positive | x | x | x | x | Later | x | Smaller | x | x | x |
| N Becker | 1990 | Branching | Stochastic | Discrete | Unnamed | Mean r and λ | Yes | No | Both | Larger | x | x | x | x | x | x | x | x | x |
| DJ Daley | 2000 | SIR-type | Stochastic | Continuous | Unnamed | Mean r and λ | Yes | Yes | Both | x | x | x | x | Earlier | x | Smaller | x | Earlier | x |
| V Andreasen | 2011 | SIR-type | Deterministic | Discrete | Unnamed | Mean r*λ | Yes | Yes | Both | x | x | x | x | x | x | Smaller | x | x | x |
| D Clancy | 2013 | SIR-type | Both | Discrete | Unnamed | Mean r*λ | Yes | Yes | Both | Smaller | x | x | x | x | x | Smaller | x | x | x |
| RI Hickson | 2014 | SIR-type | Deterministic | Continuous | Other (uniform, bimodal on [-1,1]) | Mean r*λ | Yes | Yes | Both | x | x | Smaller | x | Same | x | Smaller | x | x | x |
| RI Hickson | 2014 | SIR-type | Deterministic | Continuous | Other (uniform, bimodal on [-1,1]) | Other (median individual $R_0$) | Yes | Yes | Both | x | x | Larger | x | Earlier | x | Larger | x | x | x |
| AV Tkachenko | 2021 | Age of inf | Deterministic | Discrete | Gamma & Other (lognormal, power law) | Mean r*λ | Yes | Yes | Positive | x | x | x | x | x | x | Smaller | x | x | x |
| K Kawagoe | 2021 | SIR-type | Deterministic | Continuous | Gamma & Other (lognormal, empirical) | Mean r and λ | Yes | Yes | Positive | x | x | x | x | Earlier | x | Smaller | Larger | x | x |
| A Allard | 2023 | Network | Stochastic | Continuous | Other (Poisson) | Mean r*λ | No | Yes | Positive | x | x | x | x | x | x | x | Larger | x | x |
| Our model | | SIR-type | Stochastic | Continuous | Gamma | Mean r and λ | Yes | Yes | Both | Depends | Larger | Depends | Larger | Earlier | Earlier | Depends | Larger | Earlier | Earlier |

Table S2: The model structure, assumptions, and results for negative correlation associated with each of the 11 models included in the systematic review and our model. For model structure, the model type is either an SIR-type (SIR, SI, etc.), a branching process, an age of infection model, or a directed network. Models were set up as deterministic, stochastic, or both to explore different results. The distributions used to model the values for susceptibility and transmissibility were either discrete or continuous and the type of distribution explored in each study was a gamma distribution, an unnamed distribution where authors set their own values, a gamma distribution plus another distribution, or solely another type of distribution like a lognormal or uniform distribution. For assumptions, models compare the effects of correlation to the uncorrelated and homogeneous cases by keeping the mean susceptibility and mean transmissibility constant ("Mean $r$ and $\lambda$"), keeping the mean product of susceptibility and transmissibility constant ("Mean $r * \lambda$"), or by keeping the growth rate for the first month of the epidemic or the median product of susceptibility and transmissibility constant ("Other"). Most models assume a well-mixed population and changing susceptible population size over time. The effect of correlation is classified for each model and measure as resulting in an attribute that is smaller/earlier (blue), larger/later (orange), the same (yellow), any of these results depending on the levels of heterogeneity or value of $R_0$ (purple), or not studied (gray) in comparison to the homogeneous or no correlation case. Boxes with lighter shading indicate that the corresponding model did not explicitly conclude that effect, but it could be determined from the results presented. For negative correlation, we found that the effect of correlation on both the peak time and the time to the $j$th infection depends on the level of heterogeneity in transmission. High heterogeneity leads to an earlier peak and $j$th infection compared to the homogeneous and no correlation cases while low heterogeneity leads to these attributes being later.

| Paper Information | | Model Structure | | | | Assumptions | | | | Results for negative correlation | | | | | | | | | |
|---|---|---|---|---|---|---|---|---|---|---|---|---|---|---|---|---|---|---|---|
| | | | | | | | | | | Probability of major epidemic | | Peak size | | Peak time | | Final epidemic size | | Time to jth infection | |
| First author | Year | Model type | Deterministic? | Discrete values? | Distribution | Constant comparison | Well-mixed | Changing S | Type of correlation studied | Compare to hom | Compare to no cor | Compare to hom | Compare to no cor | Compare to hom | Compare to no cor | Compare to hom | Compare to no cor | Compare to hom | Compare to no cor |
| NS Cardell | 1989 | SIR-type | Deterministic | Continuous | Gamma | Mean r and λ | Yes | Yes | Positive | x | x | x | x | x | x | x | x | x | x |
| NS Cardell | 1989 | SIR-type | Deterministic | Continuous | Gamma | Other (growth rate for first month) | Yes | Yes | Positive | x | x | x | x | x | x | x | x | x | x |
| N Becker | 1990 | Branching | Stochastic | Discrete | Unnamed | Mean r and λ | Yes | No | Both | Smaller | x | x | x | x | x | x | x | x | x |
| DJ Daley | 2000 | SIR-type | Stochastic | Continuous | Unnamed | Mean r and λ | Yes | Yes | Both | Smaller | x | x | x | Later | x | Smaller | x | Later | x |
| V Andreasen | 2011 | SIR-type | Deterministic | Discrete | Unnamed | Mean r*λ | Yes | Yes | Both | x | x | x | x | x | x | Depends | x | x | x |
| D Clancy | 2013 | SIR-type | Both | Discrete | Unnamed | Mean r*λ | Yes | Yes | Both | x | x | x | x | x | x | Depends | x | x | x |
| RI Hickson | 2014 | SIR-type | Deterministic | Continuous | Other (uniform, bimodal on [-1,1]) | Mean r*λ | Yes | Yes | Both | x | x | Larger | x | Same | x | Larger | x | x | x |
| RI Hickson | 2014 | SIR-type | Deterministic | Continuous | Other (uniform, bimodal on [-1,1]) | Other (median individual $R_0$) | Yes | Yes | Both | x | x | Smaller | x | Later | x | Smaller | x | x | x |
| AV Tkachenko | 2021 | Age of inf | Deterministic | Discrete | Gamma & Other (lognormal, power law) | Mean r*λ | Yes | Yes | Positive | x | x | x | x | x | x | x | x | x | x |
| K Kawagoe | 2021 | SIR-type | Deterministic | Continuous | Gamma & Other (lognormal, empirical) | Mean r and λ | Yes | Yes | Positive | x | x | x | x | x | x | x | x | x | x |
| A Allard | 2023 | Network | Stochastic | Continuous | Other (Poisson) | Mean r*λ | No | Yes | Positive | x | x | x | x | x | x | x | x | x | x |
| Our model | | SIR-type | Stochastic | Continuous | Gamma | Mean r and λ | Yes | Yes | Both | Smaller | Smaller | Smaller | Smaller | Depends | Depends | Smaller | Smaller | Depends | Depends |

### 2. Peak size and final epidemic size for $R_0 = 3$

As discussed in the main text, the peak size and final epidemic size are predominantly affected by the level of heterogeneity in susceptibility with increased heterogeneity in susceptibility leading to reduced peak and final epidemic sizes. When susceptibility and transmissibility are correlated, however, positive correlations can result in larger peak and final epidemic sizes in comparison to the no correlation case while negative correlations result in smaller peaks and epidemics. Additionally, when there is a correlation between transmissibility and susceptibility, changing the magnitude of heterogeneity in transmission can impact both peak size and final epidemic size In particular, with negative correlations, increasing heterogeneity in transmission decreases peak size and final epidemic size and decreases it relative to the uncorrelated case. With positive correlations, on the other hand, increasing heterogeneity in transmission increases peak size in comparison to the uncorrelated case. This is because at a high level of heterogeneity in transmission, the individuals most likely to become infected early tend to be those that are least or most transmissible respectively.

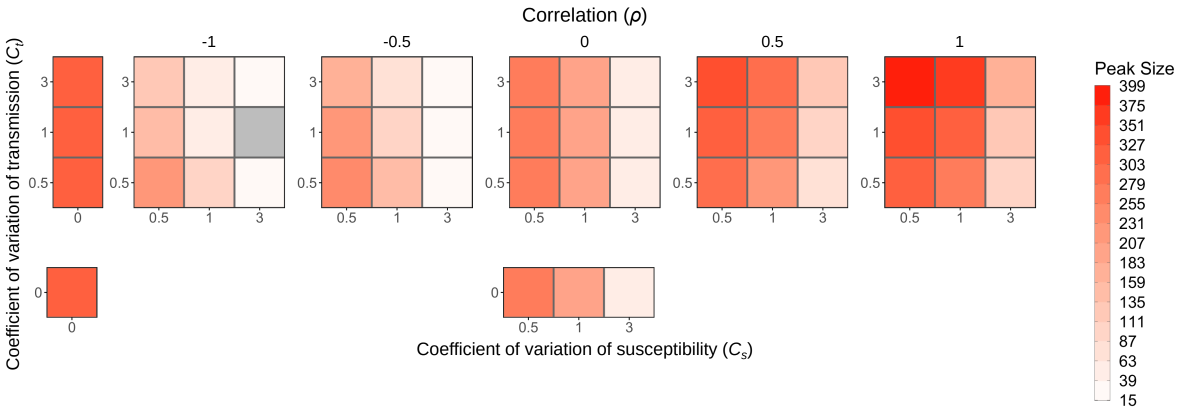

Figure S1: Peak size decreases as the level of heterogeneity in susceptibility increases, increases with positive correlation, and decreases with negative correlation. Each box is shaded to show the peak size averaged over the major epidemics from 500 simulations for various levels of heterogeneity in transmission ($C_t$), heterogeneity in susceptibility ($C_s$), and the correlation between transmissibility and susceptibility. The number of simulations that were major epidemics, which can be determined by the probability of a major epidemic in Fig 7, was different for each parameter combination, so the average in each box is based on a different sample size. The gray box represents a parameter combination that resulted in no major epidemics as defined in the text. While heterogeneity in susceptibility primarily determines the peak size, there are also effects from correlation and heterogeneity in transmission. Positive correlation results in a larger peak size, negative correlation results in a smaller peak size, and increased levels of heterogeneity in transmission enhance the effect of correlation. $N = 1000$, $I_0 = 1$, and $R_0 = 3$.

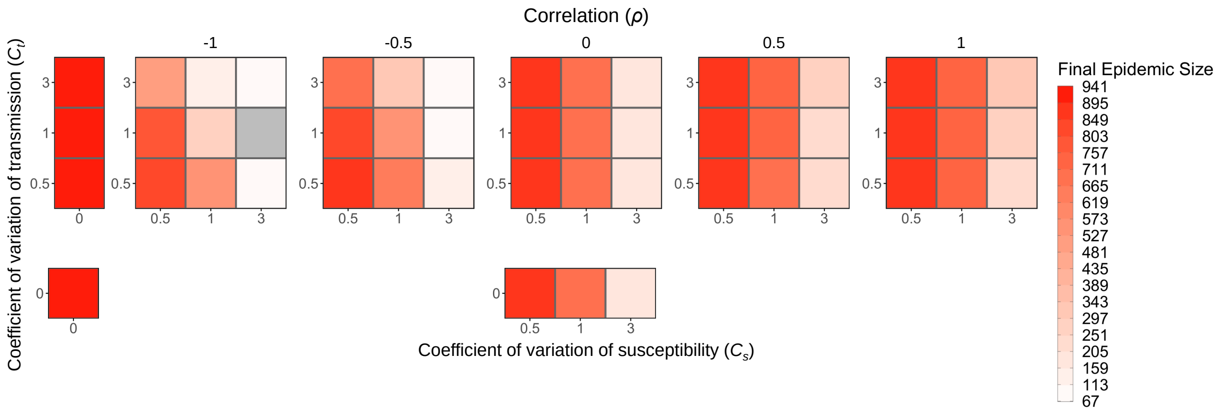

Figure S2: Final epidemic size decreases as the level of heterogeneity in susceptibility increases, increases with positive correlation, and decreases with negative correlation. Each box is shaded to show the final epidemic size averaged over the major epidemics from 500 simulations for various levels of heterogeneity in transmission ($C_t$), heterogeneity in susceptibility ($C_s$), and the correlation between transmissibility and susceptibility. The number of simulations that were major epidemics, which can be determined by the probability of a major epidemic in Fig 7, was different for each parameter combination, so the average in each box is based on a different sample size. The gray box represents a parameter combination that resulted in no major epidemics as defined in the text. While heterogeneity in susceptibility primarily determines the final epidemic size, there are also effects from correlation and heterogeneity in transmission. Positive correlation results in a larger epidemic size, negative correlation results in a smaller epidemic size, and increased levels of heterogeneity in transmission enhance the effect of correlation. $N = 1000$, $I_0 = 1$, and $R_0 = 3$.

### 3. Time to the 50th infection and time for the epidemic to reach 50% of its final size for $R_0 = 3$

In Fig S3, we simplify the data from Fig 9 in the main text, which shows the time to the $j$th infection, to show only the time to the 50th infection. We do this to more readily compare across correlations and levels of heterogeneity. Compared to the case with heterogeneity but no correlation, a positive correlation leads to the 50th infection happening earlier and a negative correlation leads to the 50th infection happening later. We also looked at the time for the epidemic to reach 50% of its final size (i.e., $j = 0.5(N - S_{\text{final}})$) to compare the speed of epidemics with different sizes and found generally similar results (Fig S4).

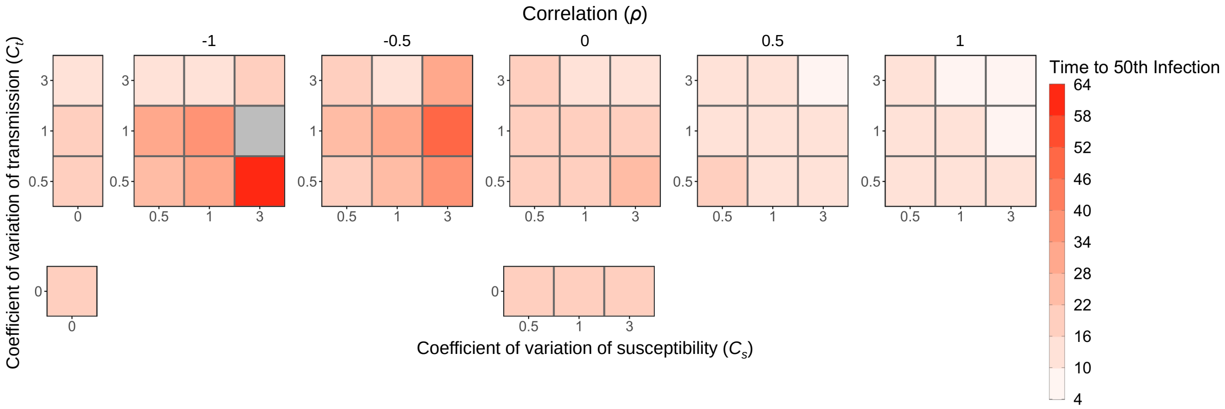

Figure S3: Time to the 50th infection is earlier with positive correlation and later with negative correlation. Each box is shaded to show the time to the 50th infection averaged over the major epidemics from 500 simulations for various levels of heterogeneity in transmission ($C_t$), heterogeneity in susceptibility ($C_s$), and the correlation between transmissibility and susceptibility. The number of simulations that were major epidemics, which can be determined by the probability of a major epidemic in Fig 7, was different for each parameter combination, so the average in each box is based on a different sample size. The gray box represents a parameter combination that resulted in no major epidemics as defined in the text. $N = 1000$, $I_0 = 1$, and $R_0 = 3$.

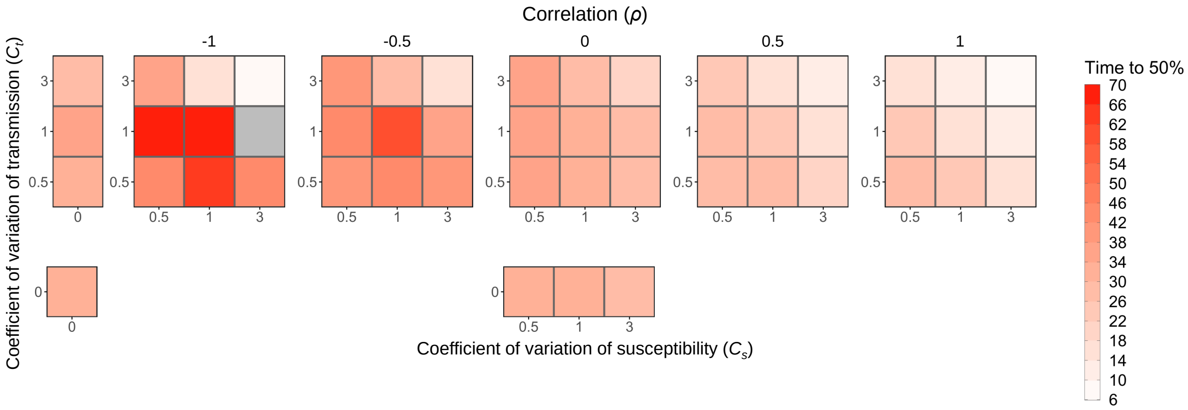

Figure S4: The time for the epidemic to reach 50% of its final size is earlier with positive correlation and later with negative correlation. Each box is shaded to show the time at which the epidemic reached 50% of its final size averaged over the major epidemics from 500 simulations for various levels of heterogeneity in transmission ($C_t$), heterogeneity in susceptibility ($C_s$), and the correlation between transmissibility and susceptibility. The number of simulations that were major epidemics, which can be determined by the probability of a major epidemic in Fig 7, was different for each parameter combination, so the average in each box is based on a different sample size. The gray box represents a parameter combination that resulted in no major epidemics as defined in the text. $N = 1000$, $I_0 = 1$, and $R_0 = 3$.

### 4. Figures for $R_0 = 1.1$

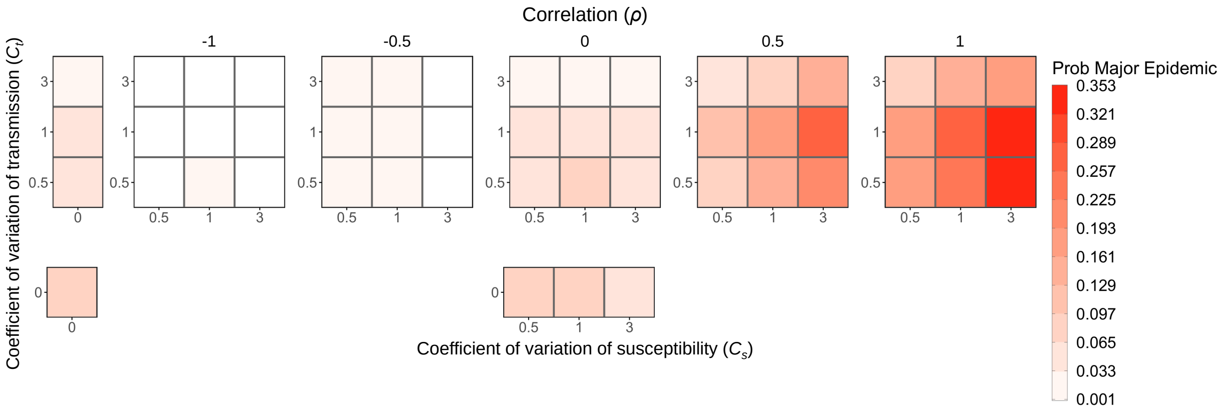

Figure S5: The probability of a major epidemic decreases as the level of heterogeneity in transmission increases, increases with positive correlation, and decreases with negative correlation. Each box is shaded to show the probability of a major epidemic, as defined in the text, averaged over the major epidemics from 500 simulations for various levels of heterogeneity in transmission ($C_t$), heterogeneity in susceptibility ($C_s$), and the correlation between transmissibility and susceptibility. Positive correlation can result in a higher probability than with no correlation or homogeneity. $N = 1000$, $I_0 = 1$, and $R_0 = 1.1$.

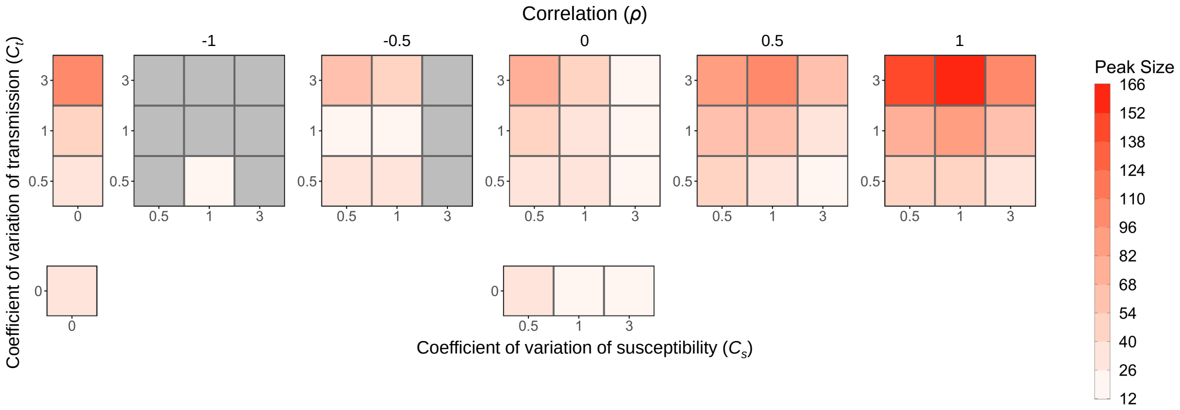

Figure S6: Peak size decreases as the level of heterogeneity in susceptibility increases and increases with positive correlation. Each box is shaded to show the peak size averaged over the major epidemics from 500 simulations for various levels of heterogeneity in transmission ($C_t$), heterogeneity in susceptibility ($C_s$), and the correlation between transmissibility and susceptibility. The number of simulations that were major epidemics, which can be determined by the probability of a major epidemic in Fig S5, was different for each parameter combination, so the average in each box is based on a different sample size. The gray boxes represent parameter combinations that resulted in no major epidemics as defined in the text. Positive correlations can result in larger peak sizes than with no correlation or homogeneity. $N = 1000$, $I_0 = 1$, and $R_0 = 1.1$.

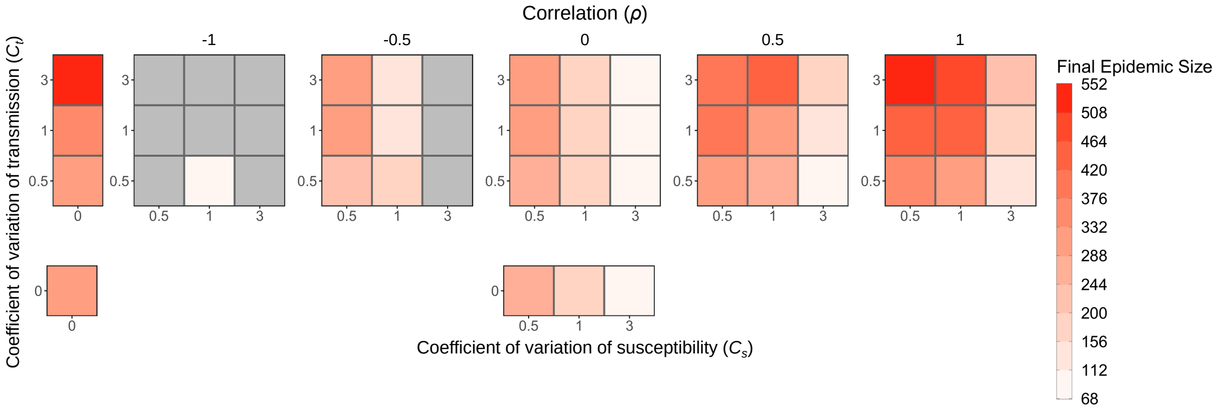

Figure S7: Final epidemic size decreases as the level of heterogeneity in susceptibility increases and increases with positive correlation. Each box is shaded to show the final epidemic size averaged over the major epidemics from 500 simulations for various levels of heterogeneity in transmission ($C_t$), heterogeneity in susceptibility ($C_s$), and the correlation between transmissibility and susceptibility. The number of simulations that were major epidemics, which can be determined by the probability of a major epidemic in Fig S5, was different for each parameter combination, so the average in each box is based on a different sample size. The gray boxes represent parameter combinations that resulted in no major epidemics as defined in the text. While heterogeneity in susceptibility has a large effect on the final epidemic size, there are also effects from correlation and heterogeneity in transmission. Positive correlation can result in a larger epidemic size than with no correlation or homogeneity, and increased levels of heterogeneity in transmission enhance this effect. $N = 1000$, $I_0 = 1$, and $R_0 = 1.1$.

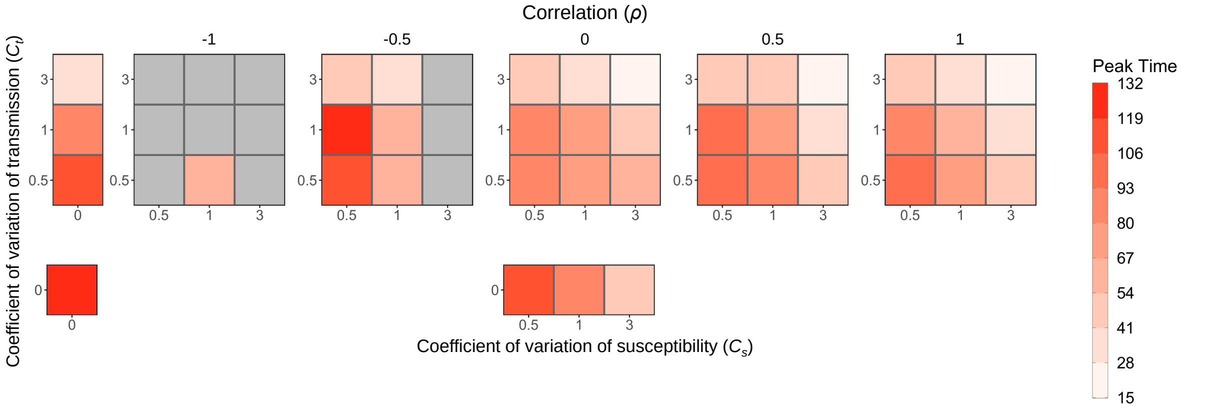

Figure S8: Peak time. Each box is shaded to show the peak time averaged over the major epidemics from 500 simulations for various levels of heterogeneity in transmission ($C_t$), heterogeneity in susceptibility ($C_s$), and the correlation between transmissibility and susceptibility. The number of simulations that were major epidemics, which can be determined by the probability of a major epidemic in Fig S5, was different for each parameter combination, so the average in each box is based on a different sample size. The gray boxes represent parameter combinations that resulted in no major epidemics as defined in the text. $N = 1000$, $I_0 = 1$, and $R_0 = 1.1$.

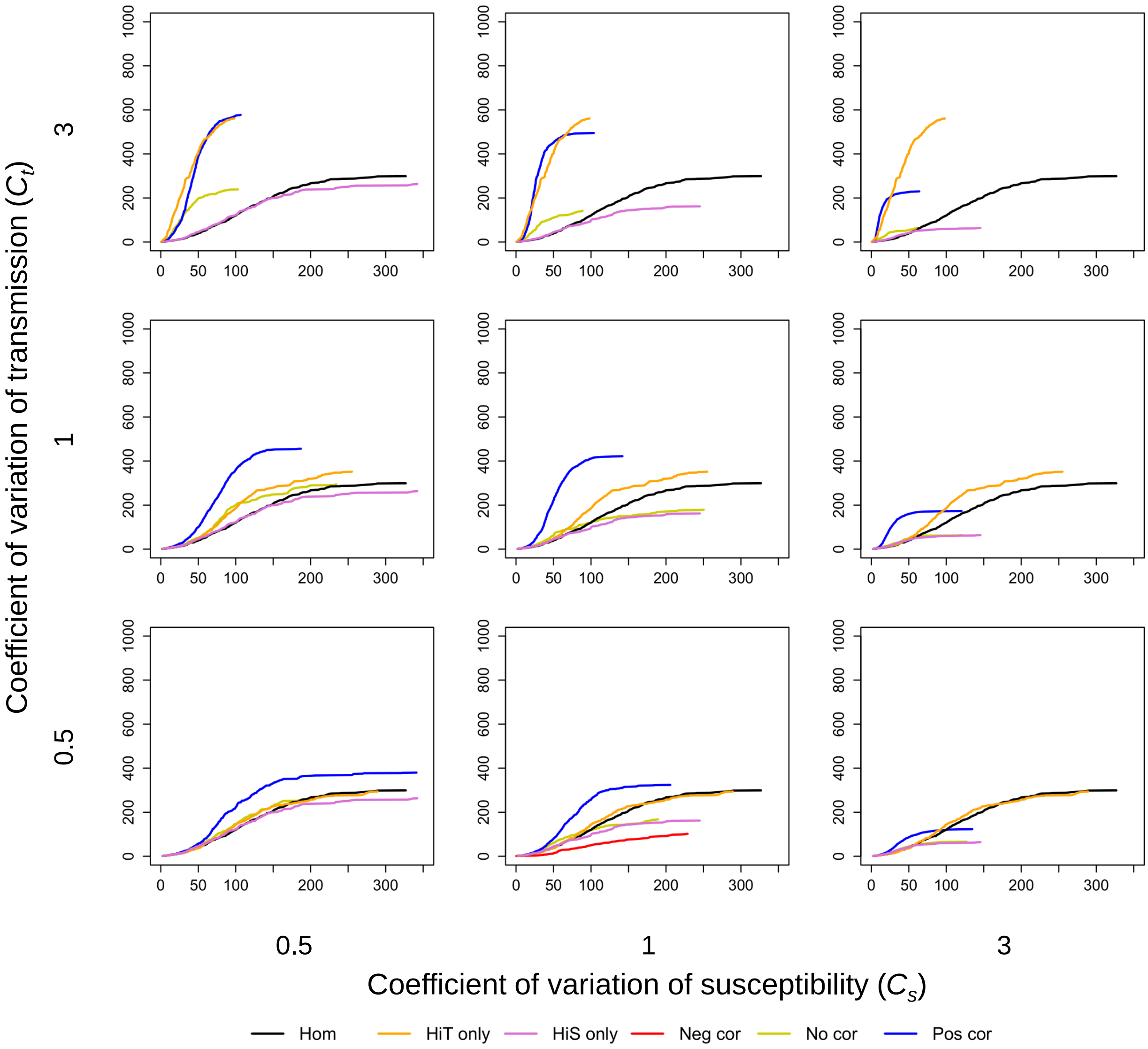

Figure S9: Time to the $j$th infection is earlier with positive correlation. The plots show the median time to the $j$th infection from the major epidemics from 500 simulations for various levels of heterogeneity in transmission ($C_t$), heterogeneity in susceptibility ($C_s$), and the correlation between transmissibility and susceptibility. Each plot includes trajectories for the cases of homogeneity (black), heterogeneity in transmission alone (orange), heterogeneity in susceptibility alone (purple), perfect negative correlation ($\rho = -1$, red), no correlation (yellow), and perfect positive correlation ($\rho = 1$, blue). The number of simulations that were major epidemics, which can be determined by the probability of a major epidemic in Fig S5, was different for each parameter combination, so each line is based on a different sample size. There are no lines for some parameter combinations because these resulted in no major epidemics as defined in the text. $N = 1000$, $I_0 = 1$, and $R_0 = 1.1$.

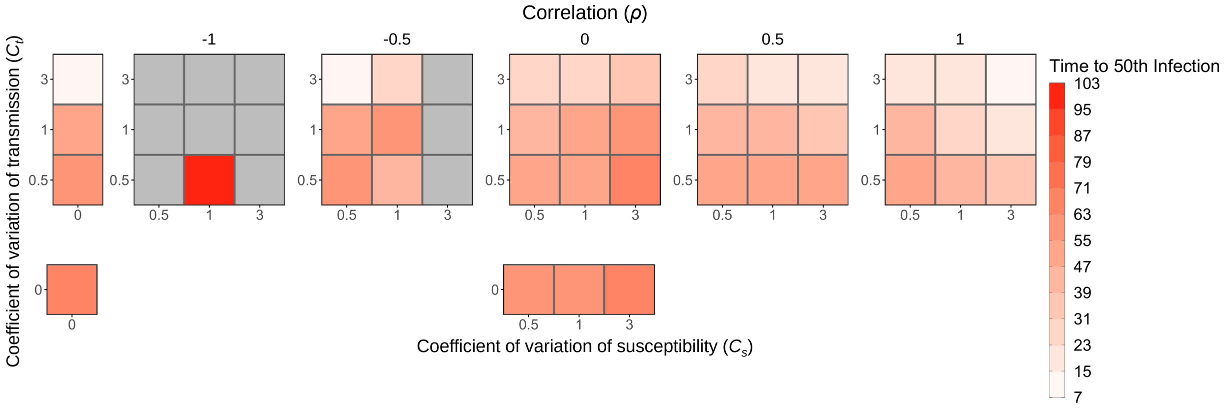

Figure S10: Time to the 50th infection. Each box is shaded to show the time to the 50th infection averaged over the major epidemics from 500 simulations for various levels of heterogeneity in transmission ($C_t$), heterogeneity in susceptibility ($C_s$), and the correlation between transmissibility and susceptibility. The number of simulations that were major epidemics, which can be determined by the probability of a major epidemic in Fig S5, was different for each parameter combination, so the average in each box is based on a different sample size. The gray boxes represent parameter combinations that resulted in no major epidemics as defined in the text. $N = 1000$, $I_0 = 1$, and $R_0 = 1.1$.

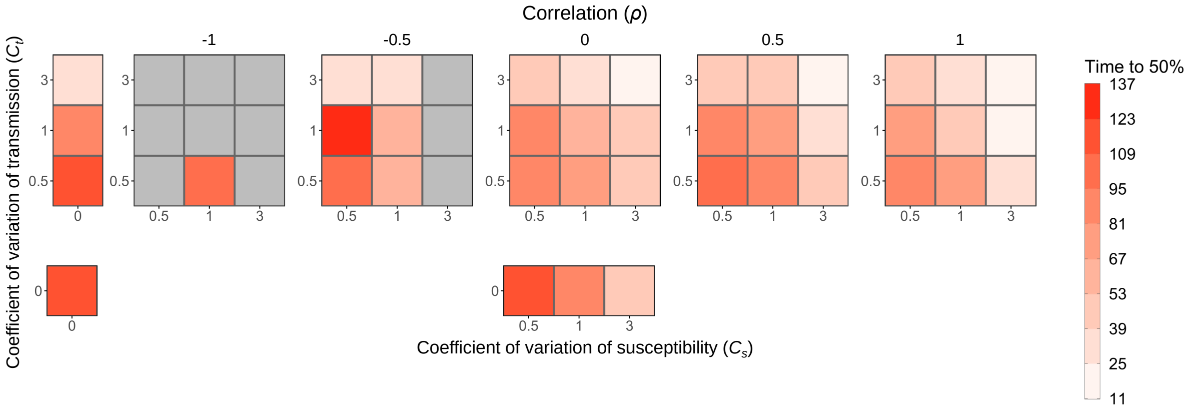

Figure S11: The time for the epidemic to reach 50% of its final size. Each box is shaded to show the time at which the epidemic reached 50% of its final size averaged over the major epidemics from 500 simulations for various levels of heterogeneity in transmission ($C_t$), heterogeneity in susceptibility ($C_s$), and the correlation between transmissibility and susceptibility. The number of simulations that were major epidemics, which can be determined by the probability of a major epidemic in Fig S5, was different for each parameter combination, so the average in each box is based on a different sample size. The gray boxes represent parameter combinations that resulted in no major epidemics as defined in the text. $N = 1000$, $I_0 = 1$, and $R_0 = 1.1$.

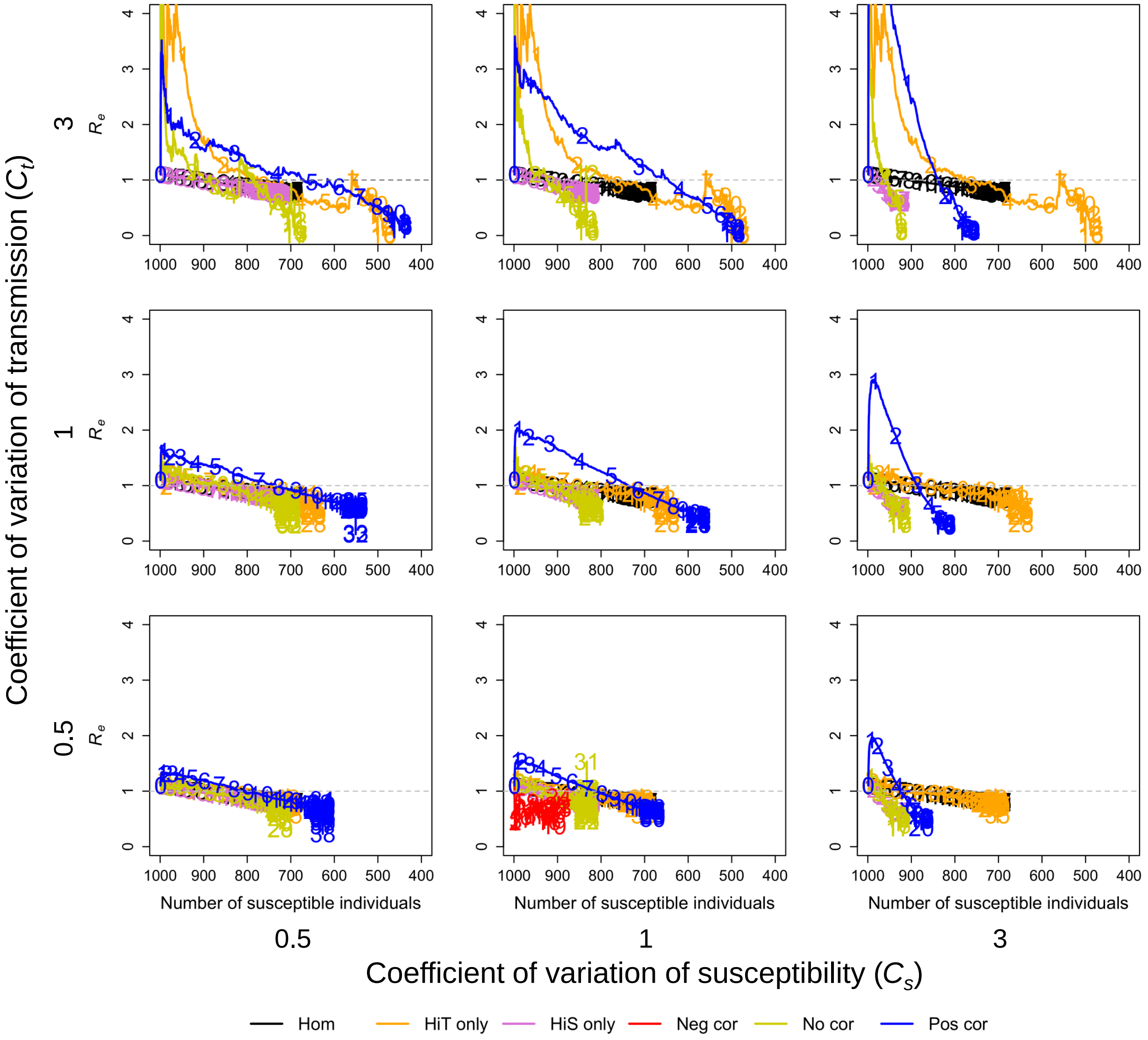

Figure S12: The effective reproductive number $R_e$ depends on both the levels of heterogeneity and the correlation. The plots show $R_e$ plotted against the number of susceptible individuals ($S$) averaged over the major epidemics from 500 simulations for various levels of heterogeneity in transmission ($C_t$), heterogeneity in susceptibility ($C_s$), and the correlation between transmissibility and susceptibility. Each plot includes trajectories for the cases of homogeneity (black), heterogeneity in transmission alone (orange), heterogeneity in susceptibility alone (purple), perfect negative correlation ($\rho = -1$, red), no correlation (yellow), and perfect positive correlation ($\rho = 1$, blue). The numbers on each trajectory represent time in the epidemic for every 10 units of time starting from the left (e.g., 1 is placed at time $t = 10$, 2 at $t = 20$, etc.). The dotted gray lines show $R_e = 1$. The number of simulations that were major epidemics, which can be determined by the probability of a major epidemic in Fig S5, was different for each parameter combination, so each line is based on a different sample size. There are no lines for some parameter combinations because these resulted in no major epidemics as defined in the text. $N = 1000$, $I_0 = 1$, and $R_0 = 1.1$.

### 5. Figures for $R_0 = 0.8$

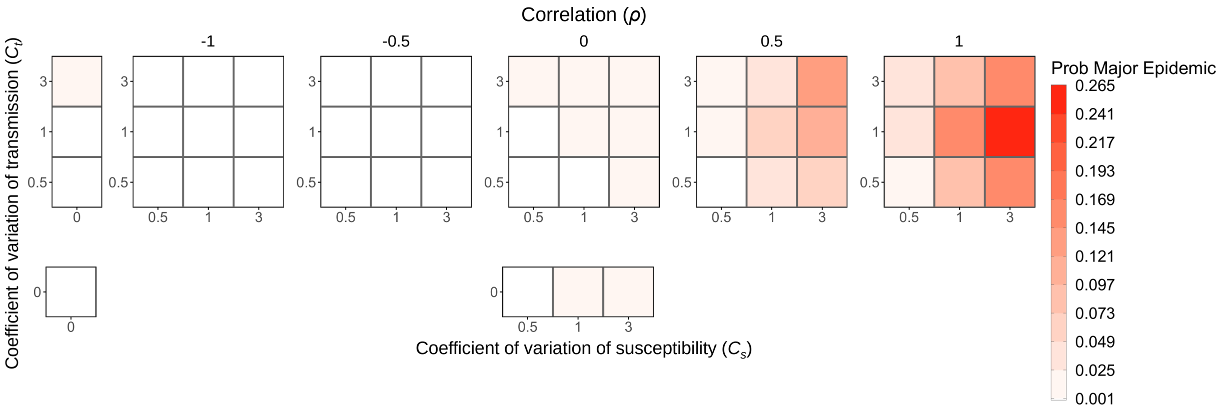

Figure S13: Major epidemics are still consistently possible with subcritical $R_0$ when there is positive correlation. Each box is shaded to show the probability of a major epidemic, as defined in the text, averaged over the major epidemics from 500 simulations for various levels of heterogeneity in transmission ($C_t$), heterogeneity in susceptibility ($C_s$), and the correlation between transmissibility and susceptibility. $N = 1000$, $I_0 = 1$, and $R_0 = 0.8$.

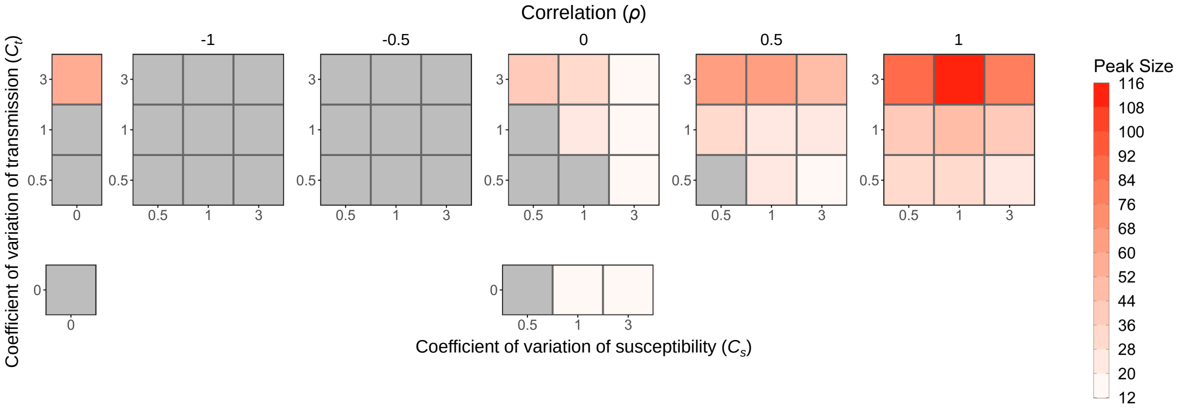

Figure S14: Peak size. Each box is shaded to show the peak size averaged over the major epidemics from 500 simulations for various levels of heterogeneity in transmission ($C_t$), heterogeneity in susceptibility ($C_s$), and the correlation between transmissibility and susceptibility. The number of simulations that were major epidemics, which can be determined by the probability of a major epidemic in Fig S13, was different for each parameter combination, so the average in each box is based on a different sample size. The gray boxes represent parameter combinations that resulted in no major epidemics as defined in the text. $N = 1000$, $I_0 = 1$, and $R_0 = 0.8$.

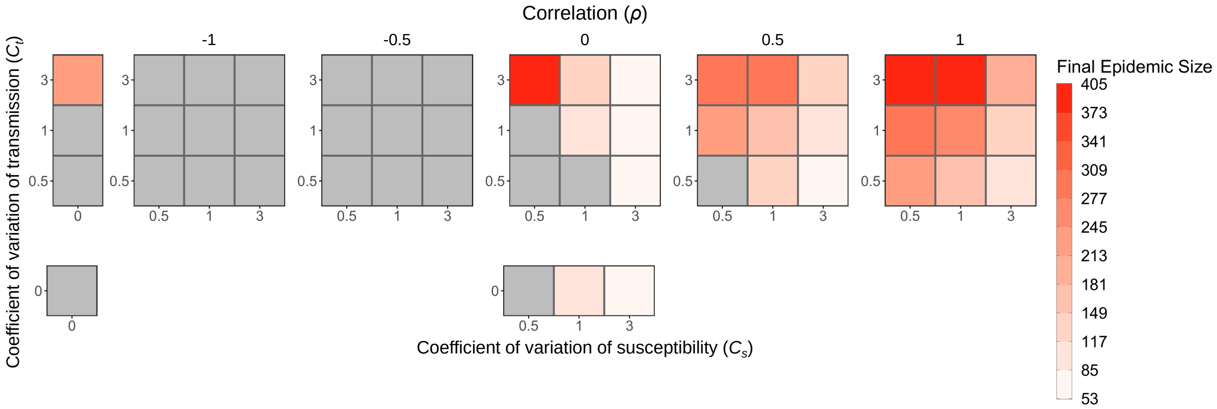

Figure S15: Final epidemic size decreases as the level of heterogeneity in susceptibility increases and increases with positive correlation. Each box is shaded to show the final epidemic size averaged over the major epidemics from 500 simulations for various levels of heterogeneity in transmission ($C_t$), heterogeneity in susceptibility ($C_s$), and the correlation between transmissibility and susceptibility. The number of simulations that were major epidemics, which can be determined by the probability of a major epidemic in Fig S13, was different for each parameter combination, so the average in each box is based on a different sample size. The gray boxes represent parameter combinations that resulted in no major epidemics as defined in the text. While heterogeneity in susceptibility has a large effect on the final epidemic size, there are also effects from correlation and heterogeneity in transmission. Positive correlation can result in large major epidemics even when none occur with no correlation or homogeneity, and increased levels of heterogeneity in transmission lead to larger epidemics. $N = 1000$, $I_0 = 1$, and $R_0 = 0.8$.

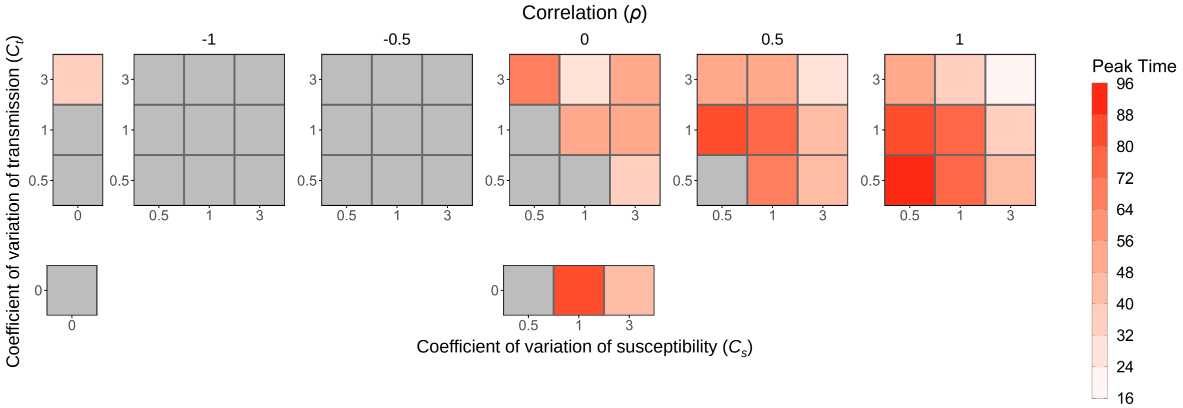

Figure S16: Peak time. Each box is shaded to show the peak time averaged over the major epidemics from 500 simulations for various levels of heterogeneity in transmission ($C_t$), heterogeneity in susceptibility ($C_s$), and the correlation between transmissibility and susceptibility. The number of simulations that were major epidemics, which can be determined by the probability of a major epidemic in Fig S13, was different for each parameter combination, so the average in each box is based on a different sample size. The gray boxes represent parameter combinations that resulted in no major epidemics as defined in the text. $N = 1000$, $I_0 = 1$, and $R_0 = 0.8$.

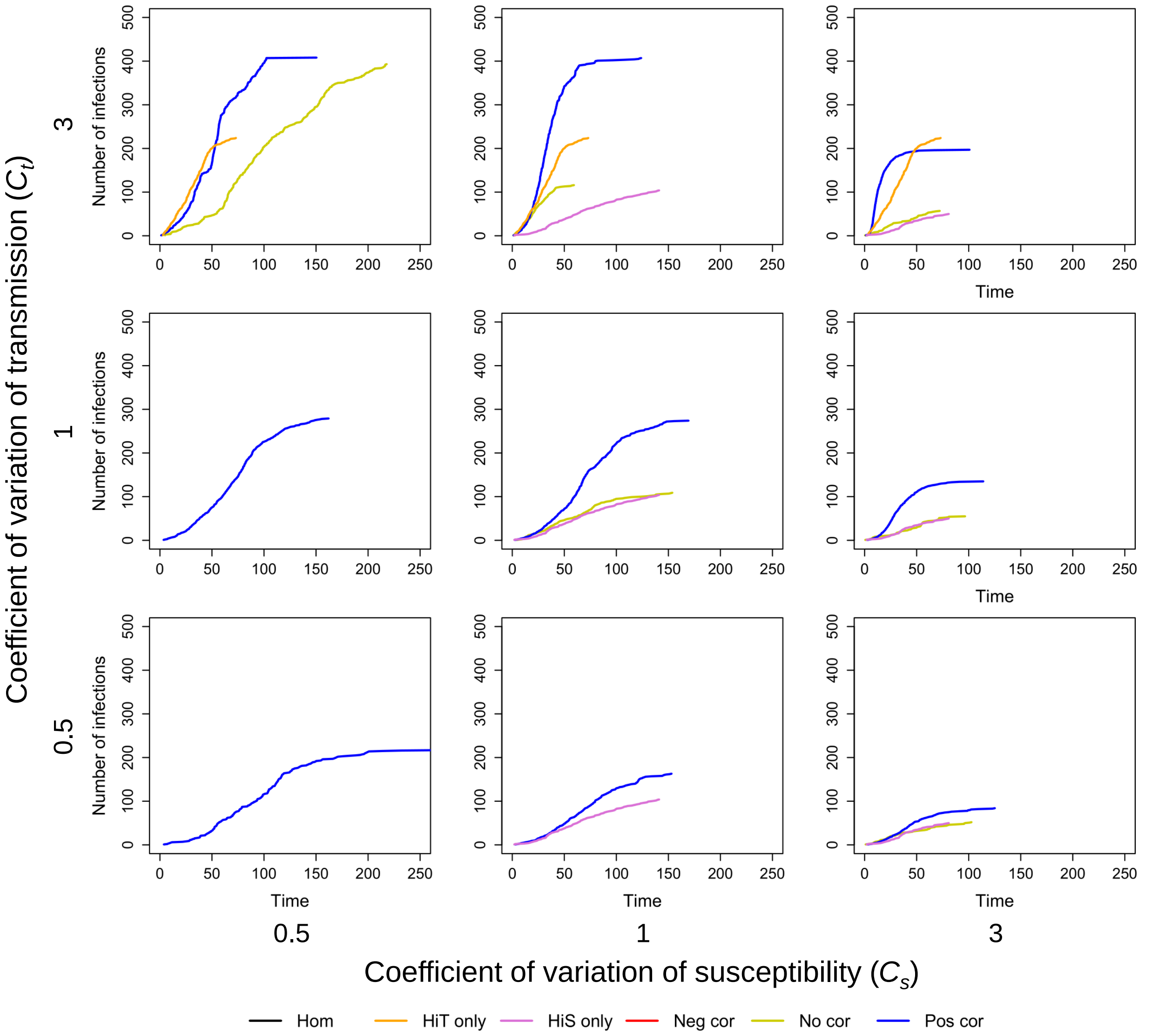

Figure S17: Time to the $j$th infection. The plots show the median time to the $j$th infection from the major epidemics from 500 simulations for various levels of heterogeneity in transmission ($C_t$), heterogeneity in susceptibility ($C_s$), and the correlation between transmissibility and susceptibility. Each plot includes trajectories for the cases of homogeneity (black), heterogeneity in transmission alone (orange), heterogeneity in susceptibility alone (purple), perfect negative correlation ($\rho = -1$, red), no correlation (yellow), and perfect positive correlation ($\rho = 1$, blue). The number of simulations that were major epidemics, which can be determined by the probability of a major epidemic in Fig S13, was different for each parameter combination, so each line is based on a different sample size. There are no lines for some parameter combinations because these resulted in no major epidemics as defined in the text. $N = 1000$, $I_0 = 1$, and $R_0 = 0.8$.

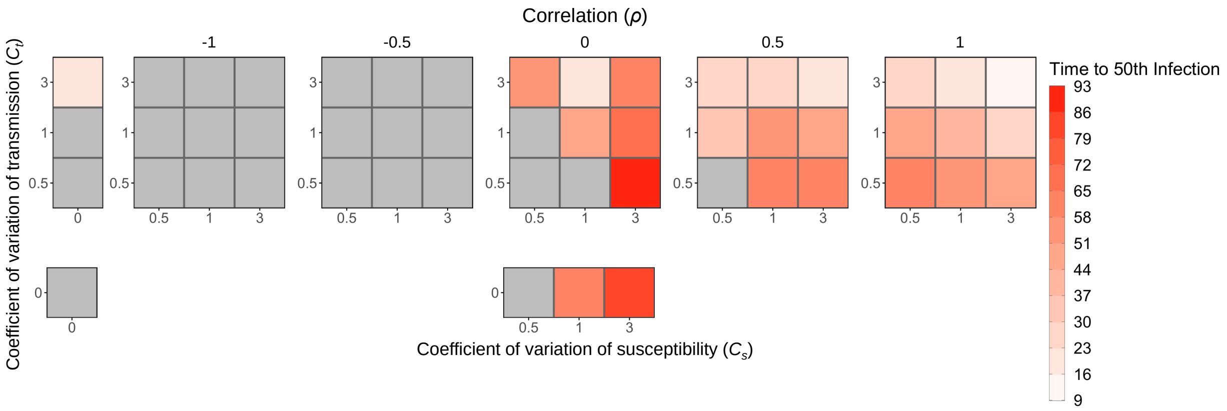

Figure S18: Time to the 50th infection. Each box is shaded to show the time to the 50th infection averaged over the major epidemics from 500 simulations for various levels of heterogeneity in transmission ($C_t$), heterogeneity in susceptibility ($C_s$), and the correlation between transmissibility and susceptibility. The number of simulations that were major epidemics, which can be determined by the probability of a major epidemic in Fig S13, was different for each parameter combination, so the average in each box is based on a different sample size. The gray boxes represent parameter combinations that resulted in no major epidemics as defined in the text. $N = 1000$, $I_0 = 1$, and $R_0 = 0.8$.

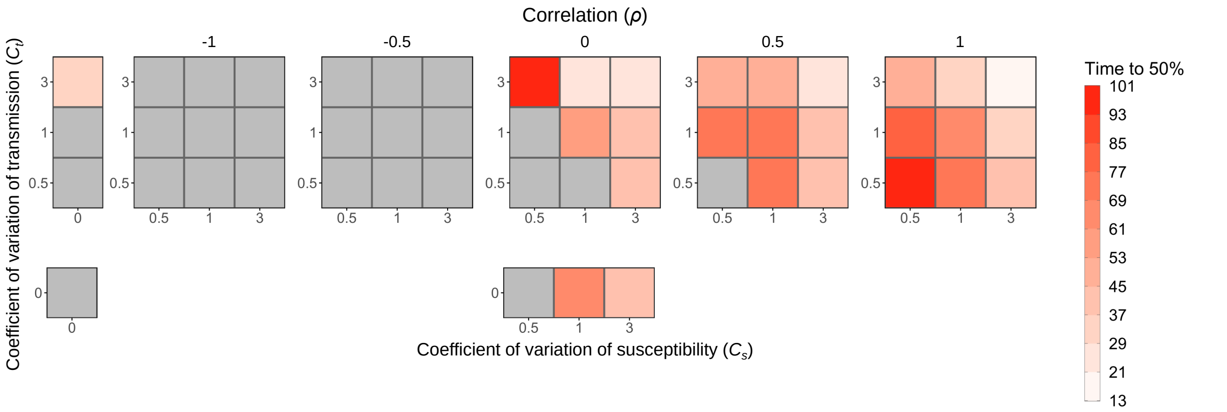

Figure S19: The time for the epidemic to reach 50% of its final size. Each box is shaded to show the time at which the epidemic reached 50% of its final size averaged over the major epidemics from 500 simulations for various levels of heterogeneity in transmission ($C_t$), heterogeneity in susceptibility ($C_s$), and the correlation between transmissibility and susceptibility. The number of simulations that were major epidemics, which can be determined by the probability of a major epidemic in Fig S13, was different for each parameter combination, so the average in each box is based on a different sample size. The gray boxes represent parameter combinations that resulted in no major epidemics as defined in the text. $N = 1000$, $I_0 = 1$, and $R_0 = 0.8$.

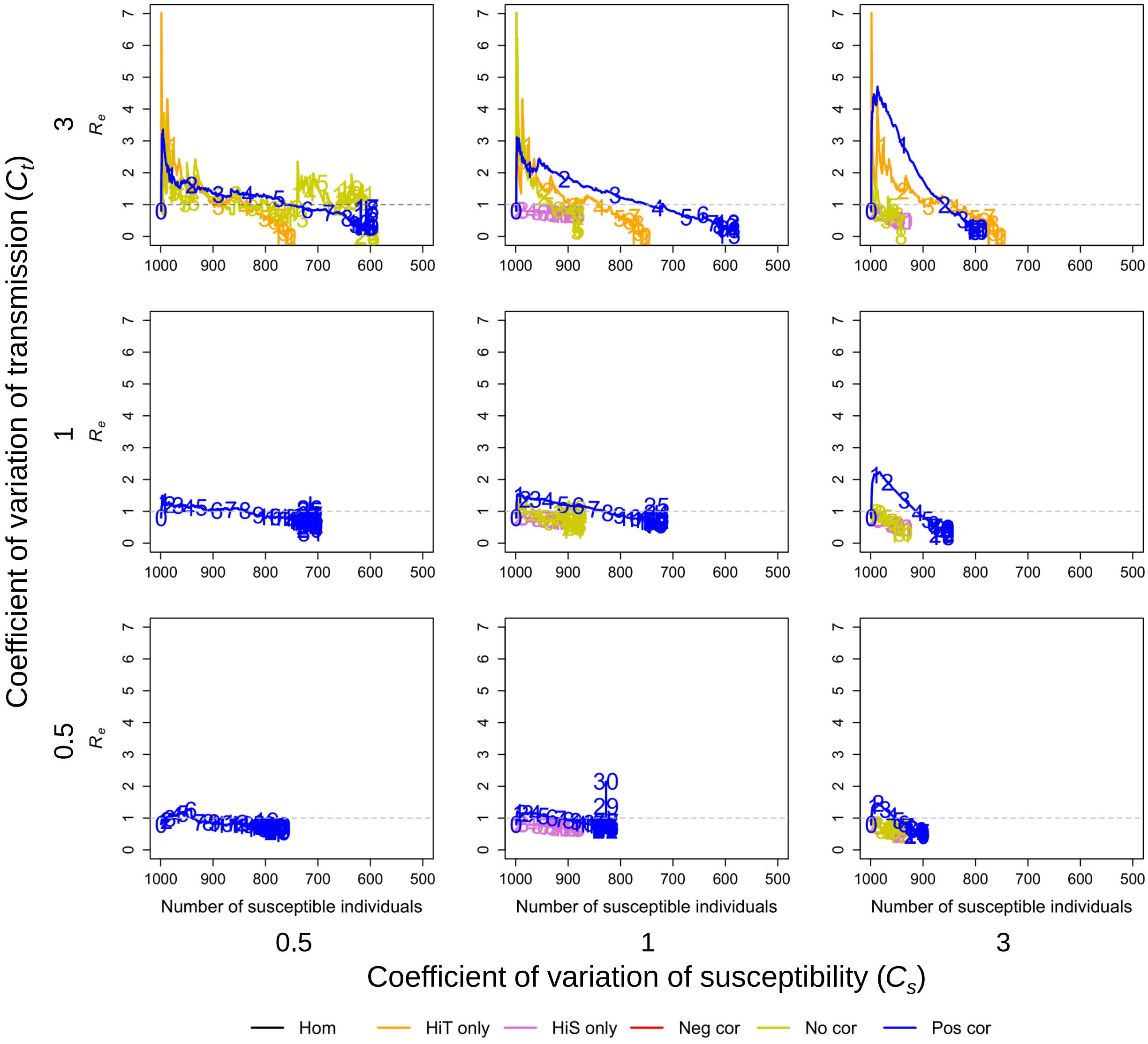

Figure S20: The effective reproductive number $R_e$. The plots show $R_e$ plotted against the number of susceptible individuals ($S$) averaged over the major epidemics from 500 simulations for various levels of heterogeneity in transmission ($C_t$), heterogeneity in susceptibility ($C_s$), and the correlation between transmissibility and susceptibility. Each plot includes trajectories for the cases of homogeneity (black), heterogeneity in transmission alone (orange), heterogeneity in susceptibility alone (purple), perfect negative correlation ($\rho = -1$, red), no correlation (yellow), and perfect positive correlation ($\rho = 1$, blue). The numbers on each trajectory represent time in the epidemic for every 10 units of time starting from the left (e.g., 1 is placed at time $t = 10$, 2 at $t = 20$, etc.). The dotted gray lines show $R_e = 1$. The number of simulations that were major epidemics, which can be determined by the probability of a major epidemic in Fig S13, was different for each parameter combination, so each line is based on a different sample size. There are no lines for some parameter combinations because these resulted in no major epidemics as defined in the text. $N = 1000$, $I_0 = 1$, and $R_0 = 0.8$.

### 6. Mpox results using $R_0$ defined as in Eq 7

We show in the main text that the 2022 mpox outbreak in New York City, as well as long term mpox dynamics, could be reasonably well explained by our model that assumes positive correlation between transmissibility and susceptibility. With zero or negative correlations, major epidemics do not occur when $R_0$ is defined using the product of mean transmissibility and mean susceptibility (Eq 6 in the main text). Here, we additionally show the effects of zero and negative correlations in this system when $R_0$ is defined using the mean product of transmissibility and susceptibility (Eq 7 in the main text). Figure S21 shows daily case counts of mpox in New York City and the trajectory of mpox as described by our SEIR model with positive (Fig S21a), zero (Fig S21b), or negative (Fig S21c) correlations between transmissibility and susceptibility under these two different definitions of $R_0$ (Eqs 6 and 7). When $R_0$ is defined by Eq 7, zero or negative correlations result in larger epidemics that overshoot the true case counts. The closer fit of the trajectory resulting from zero correlation in this case illustrates the effect of a high level of heterogeneity on disease dynamics. However, unlike the model with positive correlation, models that have no correlation or negative correlation are not able to fully capture the peak, the overall case counts, the long term persistence of mpox at low levels after the peak, and the small resurgence in case counts that occurs in 2024. This demonstrates the impact that correlations can have on disease dynamics.

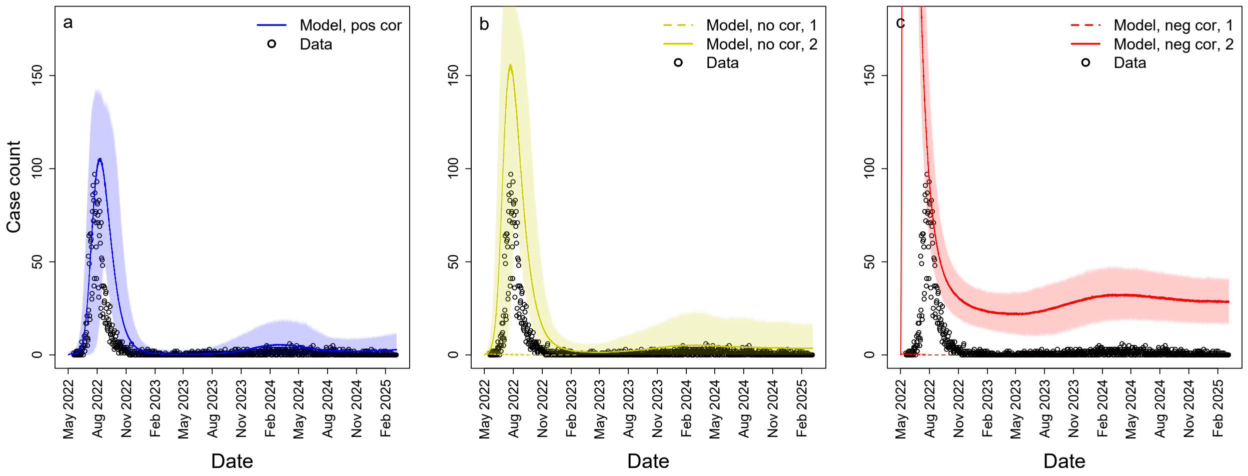

Figure S21: Mpox dynamics in New York City are generally consistent with positive correlations between transmissibility and susceptibility. Daily case counts of mpox in New York City (black circles) from May 19, 2022 to March 8, 2025 and the average number of infectious individuals from our SEIR model starting May 2, 2022 for a) positive correlation ($\rho = 1$), b) no correlation ($\rho = 0$), and c) negative correlation ($\rho = -1$). In b and c, we show the model predictions that result from defining $R_0$ as in Eq 6 (dashed, 1) versus those that result from defining $R_0$ as in Eq 7 (solid, 2). Shaded regions represent the 95% CIs of 500 simulations. The small discrepancy between our model and the daily case counts during the latter half of the 2022 epidemic is due to an observed faster than expected decline in cases, which may be attributable to vaccination and other public health measures. $C_s = C_t = 3$, $R_0 = 0.52$ (Eq 6), $R_0 = 5.2$ (Eq 7), $N = 70180$, and $E_0 = 19$.
